# Modifiable Risk Factors for Stroke, Dementia, and Late-Life Depression: A Systematic Review and DALY Weighted Risk Factors for a Composite Outcome

**DOI:** 10.1101/2024.07.08.24309905

**Authors:** Jasper R. Senff, Reinier W.P. Tack, Akashleena Mallick, Leidys Gutierrez-Martinez, Jonathan Duskin, Tamara N. Kimball, Zeina Chemali, Amy Newhouse, Christina Kourkoulis, Cyprien Rivier, Guido J Falcone, Kevin N Sheth, Ronald M Lazar, Sarah Ibrahim RN, Aleksandra Pikula, Rudolph E. Tanzi, Gregory Fricchione, H. Bart Brouwers, Gabrël J.E. Rinkel, Nirupama Yechoor, Jonathan Rosand, Christopher D. Anderson, Sanjula D. Singh

**Affiliations:** Henry and Allison McCance Center for Brain Health, Massachusetts General Hospital, Boston, MA, USA; Department of Neurology, Massachusetts General Hospital, Boston, MA, USA; Broad Institute of MIT and Harvard, Cambridge, MA, USA; Center for Genomic Medicine, Massachusetts General Hospital, Boston, MA, USA; Department of Neurology and Neurosurgery, Brain Center Rudolf Magnus, University Medical Center Utrecht, Utrecht, NL; Division of Neuropsychiatry, Massachusetts General Hospital, Boston, MA, United States; Department of Medicine, Massachusetts General Hospital, Boston, MA, United States; Department of Neurology, Yale School of Medicine, New Haven, CT, USA; Yale Center for Brain and Mind Health, Yale School of Medicine, New Haven, CT, USA; McKnight Brain Institute, Department of Neurology, School of Medicine, University of Alabama School of Medicine, Birmingham, AL, United States; Program for Health System and Technology Evaluation, Toronto General Hospital Research Institute, Department of Neurology, Toronto Western Hospital; Centre for Advancing Collaborative Healthcare & Education (CACHE), University of Toronto; Department of Medicine (Neurology), University of Toronto, Toronto, ON, Canada; Jay and Sari Sonshine Centre for Stroke Prevention and Cerebrovascular Brain Health, Krembil Brain Institute, University Health Network, Toronto, ON, Canada; Lawrence S Bloomberg Faculty of Nursing, University of Toronto, Toronto, ON, Canada; Department of Neurology, Brigham and Women’s Hospital, Boston, MA, USA

## Abstract

**Background:** At least 60% of stroke, 40% of dementia, and 35% of late-life depression (LLD) are attributable to modifiable risk factors, with great overlap due to a shared underlying pathophysiology. This study aims to systematically identify overlapping risk factors for these diseases and calculate their relative impact on a composite outcome.

**Methods:** A systematic literature review was performed in Pubmed, Embase, and PsycInfo, between January 2000 and September 2023. We included meta-analyses reporting effect sizes of modifiable risk factors on the incidence of stroke, dementia, and/or LLD. The most relevant meta-analyses were selected, and Disability Adjusted Life Year (DALY) weighted beta-coefficients were calculated for a composite outcome. The beta-coefficients were then normalized to assess relative impact.

**Results:** Our search yielded 182 meta-analyses meeting the inclusion criteria, of which 59 were selected to calculate DALY-weighted risk factors for a composite outcome. Identified risk factors included alcohol use (normalized beta-coefficient highest category: -20), blood pressure (87), BMI (42), fasting plasma glucose (57), total cholesterol (14), leisure time cognitive activity (-54), depressive symptoms (34), diet (27), hearing loss (35), kidney function (60), pain (25), physical activity (-34), purpose in life (-30), sleep (44), smoking (58), social engagement (32), and stress (32).

**Discussion:** This study identified overlapping modifiable risk factors and calculated the relative impact of these factors on the risk of a composite outcome of stroke, dementia, and LLD. These findings could guide preventative strategies and serve as an empirical foundation for future development of tools that can empower people to reduce their risk of these diseases.

**Funding:** US National Institutes of Health and American Heart Association.

## Introduction

Neurological disorders are the leading cause of disability-adjusted life years (DALYs) worldwide^1^. This is largely contributed by stroke (>143 million DALYs), dementia (>25 million DALYs), and depression (>37 million DALYs)^2^. Research indicates that at least 60% of strokes, 40% of dementia, and 35% of late-life depression (LLD) cases could be prevented or slowed down toward the limit of human life span through adequate risk factor control^3–6^. Epidemiological studies demonstrate that risk factors such as blood pressure, blood sugar, cholesterol, diet, body mass index (BMI), physical activity, smoking, and social isolation are shared among these age-related brain diseases^1,7–10^. This overlap in risk factors is, at least partially, attributable to the shared underlying pathophysiology of neurodegenerative and cerebrovascular disease, including cerebral small vessel disease (CSVD) – with multifaceted impact on cerebral circulation and brain integrity^11,12^.

Evidence-based tools, such as models or scores, can empower, educate, and motivate both patients and practitioners to facilitate behavioral changes that reduce modifiable risk factors for age-related brain diseases^13^. However, the currently available tools that address modifiable risk factors have limitations. Most existing tools for dementia, stroke, and LLD mostly focus on risk stratification of individual brain diseases ^14–23^ or together with cardiovascular disease^10^, lacking a holistic approach that addresses the shared underlying pathophysiology. In line with the recommendations of the American Heart Association (AHA) and the American Academy of Neurology (AAN)^10,24,25^, there is a need to develop, optimize, and implement novel and practical tools that address modifiable risk factors to substantiate preventive neurology in both primary care and specialized medical care worldwide.

While there are overlapping risk factors for stroke, dementia, and LLD, these factors have varying impacts on each disease^1,7–10^ To develop effective tools for addressing age-related brain diseases holistically, it is crucial to first understand how overlapping risk factors differentially impact the incidence and burden (expressed in DALYs) of these diseases, which is currently a gap in the available literature.

Therefore, this study aims to identify overlapping risk factors, obtain the most relevant effect sizes, and calculate their DALY-weighted effect on a composite outcome of stroke, dementia, and LLD.

## Methods

### Study design

The study design, registered in PROSPERO (identifier: CRD42023476939), is illustrated in Figure 1. The systematic review of the literature was conducted in line with the Joanna Briggs Institute and PRISMA guidelines^26,27^. The search design was based on a PEO (Population, Exposure, Outcome) format^28^.

**Figure 1:**
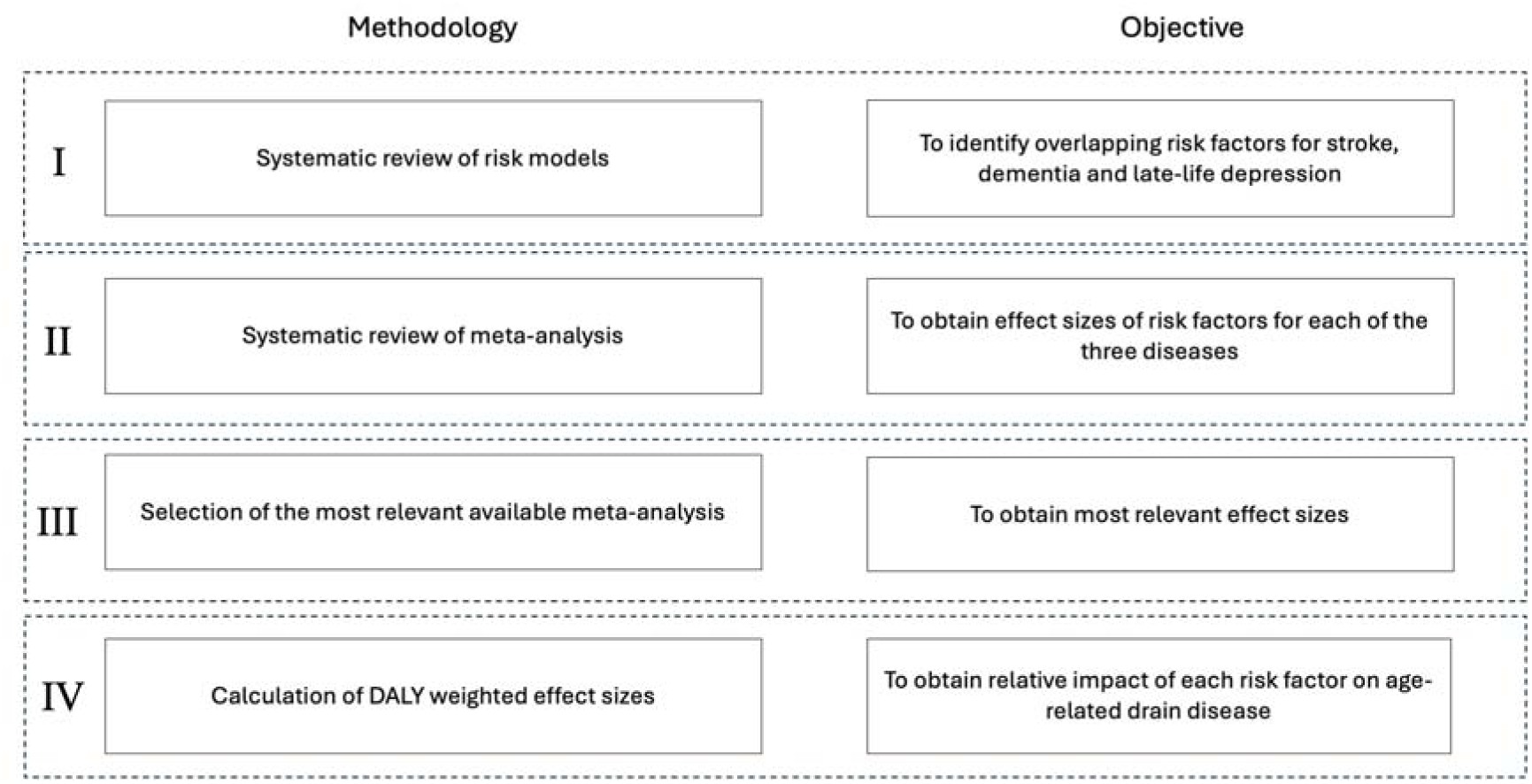
Study Design.

### Systematic review of risk models: identifying overlapping risk factors

One authors (JRS) searched PubMed and Embase from January 2000 to July 2023 (Table S1). Inclusion criteria were: (i) indexed reviews, guidelines, development, and validation studies, (ii) studies describing risk models for stroke, dementia, and/or LLD, (ii) models validated with a predictive value with a c-statistic of ≥0.70^29^, and (iii) models including modifiable risk factors. We included the most recently validated iteration of any model. Exclusion criteria were: (i) models made for a disease-specific population (e.g., stroke risk in patients with atrial fibrillation) and (ii) machine learning models^30^. Data extraction included author, publication year, model details, cohort details, statistical analysis, risk factors, and outcomes. We selected modifiable risk factors overlapping in at least two diseases for the systematic review of meta-analyses.

### Systematic review of meta-analyses: obtaining effect sizes of individual diseases per risk factor

#### Search strategy

A search was conducted in PubMed, Embase, and PsycInfo between January 2000 and September 2023, by two authors: JRS and RWPT (Table S2). Included exposures were the modifiable risk factors previously identified. The outcomes were defined as the incidence of all cause-dementia (including Alzheimer’s disease, vascular dementia, and/or all dementia), stroke (including both ischemic and hemorrhagic stroke) and/or LLD. Both authors performed title and abstract screening and full-text analyses independently from each other. Disagreements were resolved in a consensus meeting with a third reviewer (SDS). The study selection was performed using Covidence (Covidence Systematic Review Software Veritas Health Innovation, Melbourne, Australia)^31^.

#### Inclusion- and exclusion criteria

The inclusion criteria were: (i) meta-analyses of observational studies, (ii) written in English, (iii) describing a disease-risk factor relationship expressed as an effect size (Relative Risk [RR], Odds Ratio [OR] or Hazard Ratio [HR]), and (iv) risk factors defined as a dichotomized or categorical variable. Restriction to meta-analyses ensured a well-powered and feasible overview of the current literature^32^, while restriction to dichotomized or categorized ensured clinical applicability^33^. For the dietary components, we included factors as stated (either recommended or contraindicated) by the AHA and the Dietary Approaches to Stop Hypertension (DASH)^10,34^. Our exclusion criteria included: (i) disease-specific populations (e.g., patients with atrial fibrillation) (ii) treatments (e.g., cholesterol levels in statin treatment), and (iii) composite outcomes (e.g., cardiovascular disease instead of stroke) or subtypes of the outcomes (e.g., ischemic stroke only).

#### Data extraction

Data extract was performed by one of two authors, JRS and RWPT. Extracted data included first author, year of publication, exposure definition, outcome definition, effect size including 95% confidence interval (CI), number of included studies in meta-analyses, total sample size, number of outcomes, level of heterogeneity (I^2^), risk of bias (ROB) tool (e.g., Newcastle-Ottawa Scale), ROB assessment, and publication bias (Egger’s test, Begg’s test, funnel plot).

#### Study selection to identify the most relevant effect sizes

Study selection to identify the most relevant effect sizes was performed by two authors (JRS and RWPT) individually. Disagreements were resolved in a consensus meeting. For each factor, the most recent meta-analysis was included. Exceptions were made if an earlier meta-analysis had a sample size at least 20% larger. If two meta-analysis had similar sample size selection was based on a difference in quality assessed through heterogeneity level, ROB assessment, and publication bias. The initial selection was based on studies that reported RR. If no studies reporting RR were available, studies reporting HR, or if not available OR, were selected.

### DALY weighted risk factors for a composite outcome

#### Risk factor definition

Risk factors cut-offs aligned with the AHA guidelines and Life’s Essential 8 where available^10^.

### Statistical Analysis

Table S4 shows the statistical methodology. To standardize effect sizes and corresponding 95% confidence interval (CI) we transformed HR and OR to RR based on the disease-specific incidence rates (r)^35^. We obtained disease-specific DALYs using the most recent incidence rates from the Global Burden of Disease study 2019 : “Stroke” (157.99 per 100,000), “Alzheimer diseases and other related dementias” (93.52 per 100,000), and “Major Depressive Disorder” (3551.60 per 100,000)^2^. To calculate weighted effect size for a composite outcome of stroke, dementia, and LLD, the relative risks with corresponding 95% CI for each risk factor category were weighted according to their attributed burden, expressed in DALYs (stroke: 5.654%, dementia: 0.997%, LLD: 1.461%)^2^. If there was no effect size for a certain disease-risk factor relationship, the disease was not included in the weighting. The relative risks that remained significant after calculating the composite effect size were log transferred into beta (p) coefficients. To enhance interpretation and assess the relative impact of the risk factors on a composite outcome, we normalized the beta-coefficient. The lowest /’5-coefficient was normalized to 1, scaling all other /’5-coefficients by the same factor^15^.

## Results

### Systematic review of risk models: Identification of overlapping modifiable risk factors

In total, 37 articles describing 54 risk models met inclusion criteria (Figure S1). Of the models, 36 (67%) were on stroke, 16 (30%) on dementia, and two (4%) for LLD; none addressed a composite outcome. Table S4 provides an overview of the included models.. The models identified 18 modifiable factors that overlapped in at least two outcomes: (1) alcohol consumption, (2) blood pressure, (3) body mass index (BMI), (4) blood sugar, (5) cholesterol, (6) cognitive activity, (7) depressive symptoms, (8) diet, (9) hearing impairment, (10) kidney function, (11) pain, (12) physical activity, (13) self-rated health, (14) sense of belonging, (15) sleep, (16) smoking, (17) social engagement, and (18) stress.

### Systematic review of meta-analyses: obtaining effect sizes of individual diseases per risk factor

Subsequently, our systematic review of meta-analyses yielded 182 articles that met our predefined criteria, encompassing 426 effect sizes (stroke N=260 [61%], dementia N=157 [37%], LLD N=9 [2%]) on 17 modifiable risk factors (Figure 2). Reported effect size metrics were RRs (N=280 [66%]), HRs (N=111 [26%]) and ORs (N=35 [8.2%]). All studies that met our criteria are publicly available at https://www.zotero.org/groups/5402286/sroma/collections/PY6XGESM.

**Figure 2:**
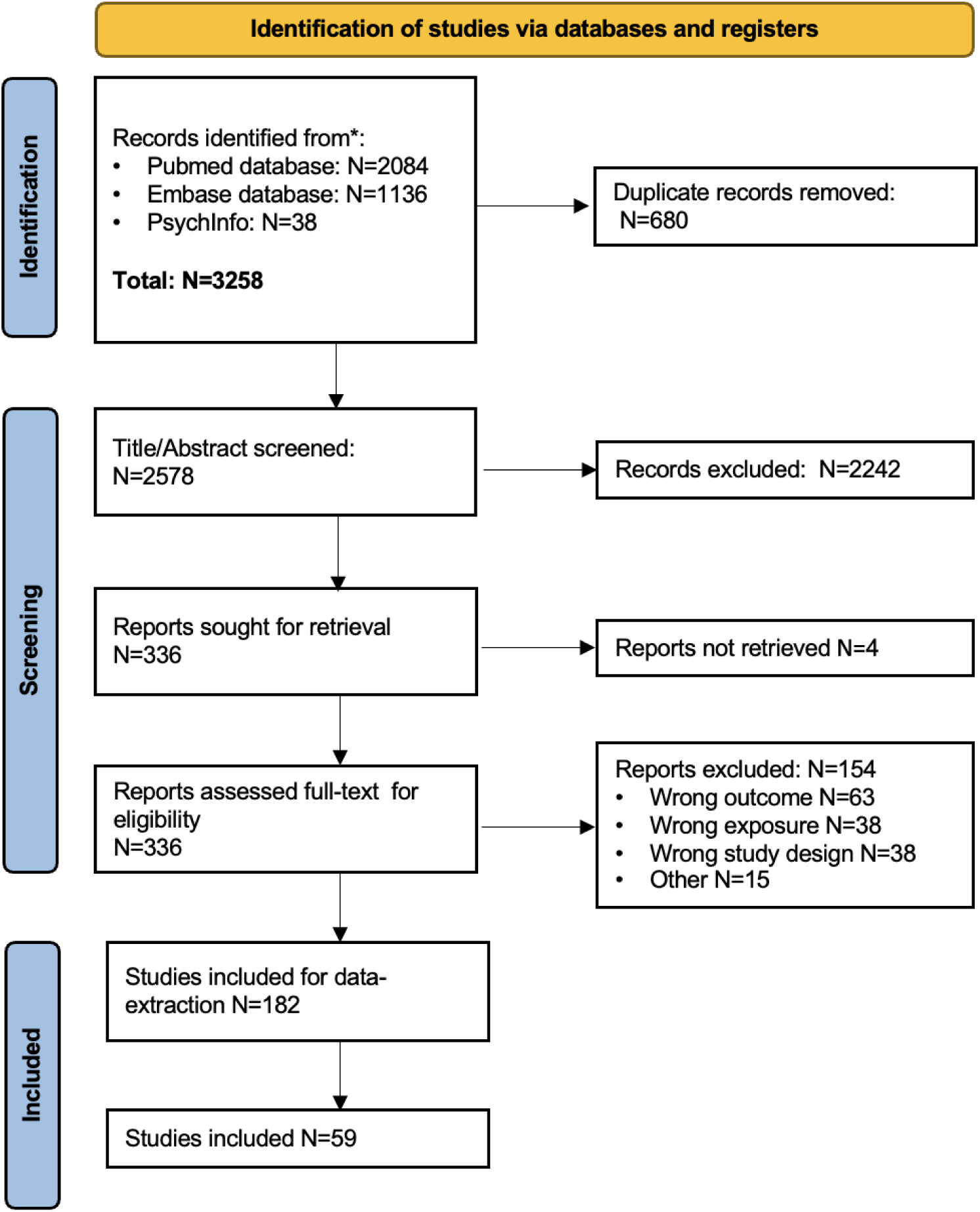
Prisma flowchart of the systematic review of meta-analyses.

#### Overview of meta-analyses

##### Alcohol

Seven meta-analyses described alcohol intake as a risk factor. For stroke incidence, effect sizes ranged from 0.80 (95%CI:0.72-0.90) – 0.89 (95%CI:0.76-1.06) for low alcohol intake (<15 gr/day), 0.79 (95%CI:0.69-0.91) – 1.10 (95%CI:0.97-1.24) for moderate alcohol intake (15-30 gr/day), and 1.19 (95%CI:0.93-1.52) – 1.64 (95%CI:1.39-1.93) for heavy alcohol intake (>30 gr/day) (reference no alcohol intake)^36–40^. For dementia incidence, effect sizes ranged from 0.74 (95%CI:0.61-0.91) – 0.75 (95%CI:0.51-1.11) for low-, 0.58 (95%CI:0.38-0.90) – 0.74 (95%CI:0.61-0.91) for moderate-, and 1.00 (95%CI:0.39-2.59) – 1.84 (95%CI:1.01-3.34) for heavy alcohol intake^41,42^. No meta-analyses were found that assessed the relationship between alcohol intake and LLD incidence.

##### Blood Pressure

Twelve meta-analyses described blood pressure as a risk factor. For stroke incidence, effect sizes ranged from 1.22 (95%CI:0.95-1.57) – 1.66 (95%CI:1.51-1.81) for low pre-hypertension (120-129/80-84mmHg), 1.79 (95%CI:1.49-2.16) – 1.95 (95%CI:1.73-2.21) for high pre-hypertension (130-139/85-80 mmHg) and 2.23 (95%CI:2.01-2.48) – 10.92 (95%CI:7.07-16.86) for hypertension (>140/90 mmHg) (reference blood pressure < 120/80 mmHg)^36,43–48^. For dementia incidence, effect sizes for hypertension ranged from 0.98 (95%CI:0.72-1.33) – 1.41 (95%CI:1.23-1.62)^49–52^. No meta-analyses were found that assessed the relationship blood pressure and LLD incidence.

##### Body Mass Index

Fifteen meta-analyses described BMI as a risk factor. For stroke incidence, the effect size for underweight (<18.5 kg/m^2^) was 0.93 (95%CI:0.82-1.06), for overweight (25-29.9 kg/m^2^) it ranged from 1.18 (95%CI:0.98-1.42) - 2.01 (95%CI:1.65-2.47), and for obesity (≥30kg/m^2^) it ranged from 1.16 (95%CI:1.01-1.35) - 1.47 (95%CI:1.02-2.11) (reference BMI ranging 18.5-25kg/m^2^)^36,47,48,53,54^.

For dementia incidence, the effect sizes for underweights ranged from 0.92 (95%CI:0.74-0.92) - 1.42 (95%CI:1.12-1.80), for overweight from 0.82 (95%CI:0.74-0.92) - 1.26 (95%CI:1.1-1.44), and for obesity from 0.78 (95%CI:0.70-0.86) - 1.79 (95%CI:1.31-2.41)^50,51,55–62^. No meta-analyses were found that assessed the relationship BMI and LLD incidence.

##### Blood Sugar

Six meta-analyses described blood sugar as a risk factor. For stroke incidence, effect sizes ranged from 1.08 (95%CI:0.94-1.23) - 1.21 (95%:1.02-1.44) for prediabetic blood sugar levels (fasting plasma glucose [FPG] 100-126 mg/dL), and from 1.79 (95%CI:1.68-1.91) -2.15 (95%CI:1.76-2.63) for diabetic blood sugar levels (FPG >126 mg/dL) (reference FPG <100 mg/dL)^36,63–65^. For dementia incidence, effect sizes ranged from 0.72 (95%CI:0.56-0.92) -1.22 (95%CI:1.06-1.41) for prediabetic blood sugar levels, and from 1.21 (95%CI:1.06-1.37) - 1.49 (95%CI:1.10-2.03) for diabetic blood sugar levels^51,66^. No meta-analyses were found that assessed the relationship blood sugar and LLD incidence.

##### Cholesterol

Fifteen meta-analyses described cholesterol as a risk factor. For stroke incidence, effect sizes ranged from 0.99 (95%CI:0.87-1.12) – 1.14 (95%CI:1.03-1.27) for high total cholesterol, and was 1.09 (95%CI:0.85-1.39) for low-density lipoprotein (LDL) (reference lowest quartiles cholesterol level)^36,43,47,48,67–71^. For dementia incidence, effect sizes ranged from 1.03 (95%CI:0.74-1.43) – 1.82 (95%CI:1.27-2.6) for high total cholesterol^50,51,56,72,73^. For LLD, the effect size for the presence of dyslipidemia was 1.08 (95%CI:0.91-1.28)^74^.

##### Cognitive activity

One meta-analysis described cognitive activity as a risk factor for dementia incidence, with an effect size of 0.61 (95%CI:0.42-0.90) (reference no cognitive activity)^75^. No meta-analyses were found that assessed the relationship cognitive activity and stroke or LLD incidence.

##### Depressive symptoms

One meta-analysis described depressive symptoms as a risk factor for stroke incidence, with an effect size of 1.36 (95%CI1.13-1.51) (reference no depressive symptoms)^76^. No meta-analyses were found that assessed the relationship depressive symptoms and dementia or LLD incidence.

##### Diet

For stroke, a meta-analysis was included for all 11 dietary components^77–87^. For dementia, meta-analyses for dairy, fish, sugar-sweetened beverages and saturated fats intake were retrieved and included^88–91^. No meta-analyses were found that assessed the relationship diet and LLD incidence.

##### Hearing loss

Three meta-analyses described hearing loss as an risk factor. The effect size was 1.33 (95%CI:1.18-1.49) for stroke^92^, 1.59 (95%CI:1,37-1.86) for dementia^93^, and 1.47 (95%CI:1.31-1.65) for LLD incidence (reference no hearing loss)^94^.

##### Kidney Function

Four meta-analyses described kidney function as a risk factor. For stroke incidence, the effect size ranged from 1.07 (95%CI:0.98-1.17) - 1.10 (95%CI:1.03-1.19) for mild-, was 1.43 (95%CI:1.33-1.54) for moderate-(eGFR 30-60mL/min/1.73m^2^) and 1.70 (95%CI:1.47-1.96) for severe kidney disease (eGFR <30mL/min/1.73m^2^) (reference eGFR ≥90mL/min/1.73min2)^36,95,96^. For dementia incidence, the effect size was 1.14 (95%CI:1.06-1.22) for mild-, 1.31 (95%CI:0.92-1.87) for moderate-, and 1.91 (95%CI:1.21-3.01) for severe kidney disease^97^. No meta-analyses were found that assessed the relationship depressive symptoms and LLD incidence.

##### Pain

One meta-analysis described the pain as a risk factor for dementia incidence, with an effect size of 1.26 (95%CI:1.18-1.35) (reference no pain)^98^. No meta-analyses were found that assessed the relationship pain and stroke or LLD incidence.

##### Physical activity

Eleven meta-analyses described physical activity as a risk factor. For stroke incidence, effects sizes for a moderate level of physical activity ranged from 0.64 (95%CI:0.48-.0.87) – 0.85 (95%CI:0.78-0.93), and was 0.73 (0.67-0.79) for a high level of physical activity (reference low level of physical activity) ^43,99–101^. For dementia incidence, effect sizes for a moderate level of physical activity ranged from 0.76 (95%CI:0.61-0.94) – 0.80 (95%CI:0.67-0.94), and from 0.63 (95%CI:0.45-0.89) – 0.80 (95%CI:0.77-0.84) for a high level of physical activity^51,101–106^. No meta-analyses were found that assessed the relationship physical activity and LLD incidence.

##### Purpose in life

Three meta-analyses described purpose in life as a risk factor for dementia^107–109^, with effect sizes ranging from 0.76 (95%CI:0.72-0.79) – 0.81 (95%CI:0.78-0.85) (reference no purpose in life)^107–109^. No meta-analyses were found that assessed the relationship purpose in life and stroke or LLD incidence.

##### Sleep

Fourteen meta-analyses described sleep as a risk factor. For stroke incidence, the effect sizes of short sleep (≤6 hours) ranged from 1.00 (95%CI:0.97-1.24) – 1.71(95%CI:1.39-2.02), for long sleep (≥8 hours) it ranged from 1.12 (95%CI:1.01-1.24) – 2.12 (95%CI:1.51-2.73), and for insomnia the effect size was 1.55 (95%CI:1.39-1.72) (reference 6-8 hours of sleep)^36,110–117^. For dementia incidence, the effect size for long sleep was 1.77 (95%CI:1.32-2.37), for short sleep 1.20 (95%CI:0.91-1.59), for insomnia it ranged 1.17 (95%CI 0.95-1.43) – 1.53 (95%CI:1.07-2.18), and for sleep disturbance, it was 1.19 (95%CI1.11-1.29)^118–120^. For LLD, the effect sizes for sleep disturbance ranged from 1.2 (95%CI:0.80-1.70) – 1.82 (95%CI:1.69-1.97)^121,122^.

##### Smoking

Fifteen meta-analyses described smoking as a risk factor. For stroke incidence, the effect sizes ranged from 1.08 (95%CI:1.03-1.13) – 1.30 (95%CI:0.93-1.81) for former smokers, and from 1.31 (95%CI:1.20-1.43) – 1.84 (1.72-.198) for current smokers (reference never smoked)^36,43,47,48,123–126^. For dementia incidence, the effect sizes ranged from 0.99 (95%CI:0.81-1.21) – 1.01 (95%CI:0.96-1.06) for former smokers, and from 1.27 (95%CI:1.02-1.60) – 1.30 (95%CI:1.18-1.45) for current smokers ^50,51,56,127–129^. For LLD, the effect size for current smoking was 1.35 (95%CI:1.00-1.81)^74^.

##### Social Engagement

Seven meta-analyses described social engagement as a risk factor. For stroke incidence, the effect size for social isolation/loneliness was 1.32 (95%CI:1.04-1.68), and 0.77 (95%CI: 0.57-1.04) for having a larger social network (reference not being lonely / small social network)^130,131^. For dementia incidence, the effect size for social isolation/loneliness ranged from 1.23 (95%CI:1.16-1.31) – 1.58 (95%CI0.80-3.12), and was 0.81 (95%CI:0.74-0.89) for having a larger social network^132–136^. No meta-analyses were found that assessed the relationship social engagement and LLD incidence.

##### Stress

Two meta-analyses described stress as a risk factor. For stroke incidence, the effect size was 1.33 (95%CI:1.17-1.50) (reference no perceived stress)^137^. For dementia incidence, the effect size was 1.44 (95%CI:1.07-1.95)^138^. No meta-analyses were found that assessed the relationship stress and LLD incidence.

### Study selection to identify the most relevant effect sizes

Of 182 meta-analyses that met the inclusion criteria, 59 were selected to calculate DALY-weighted risk factors for a composite outcome. An overview of the selected studies is presented in Table 1, with the reasoning for selection detailed in Table S5. Study characteristics are reported in Table S6.

**Table 1:**
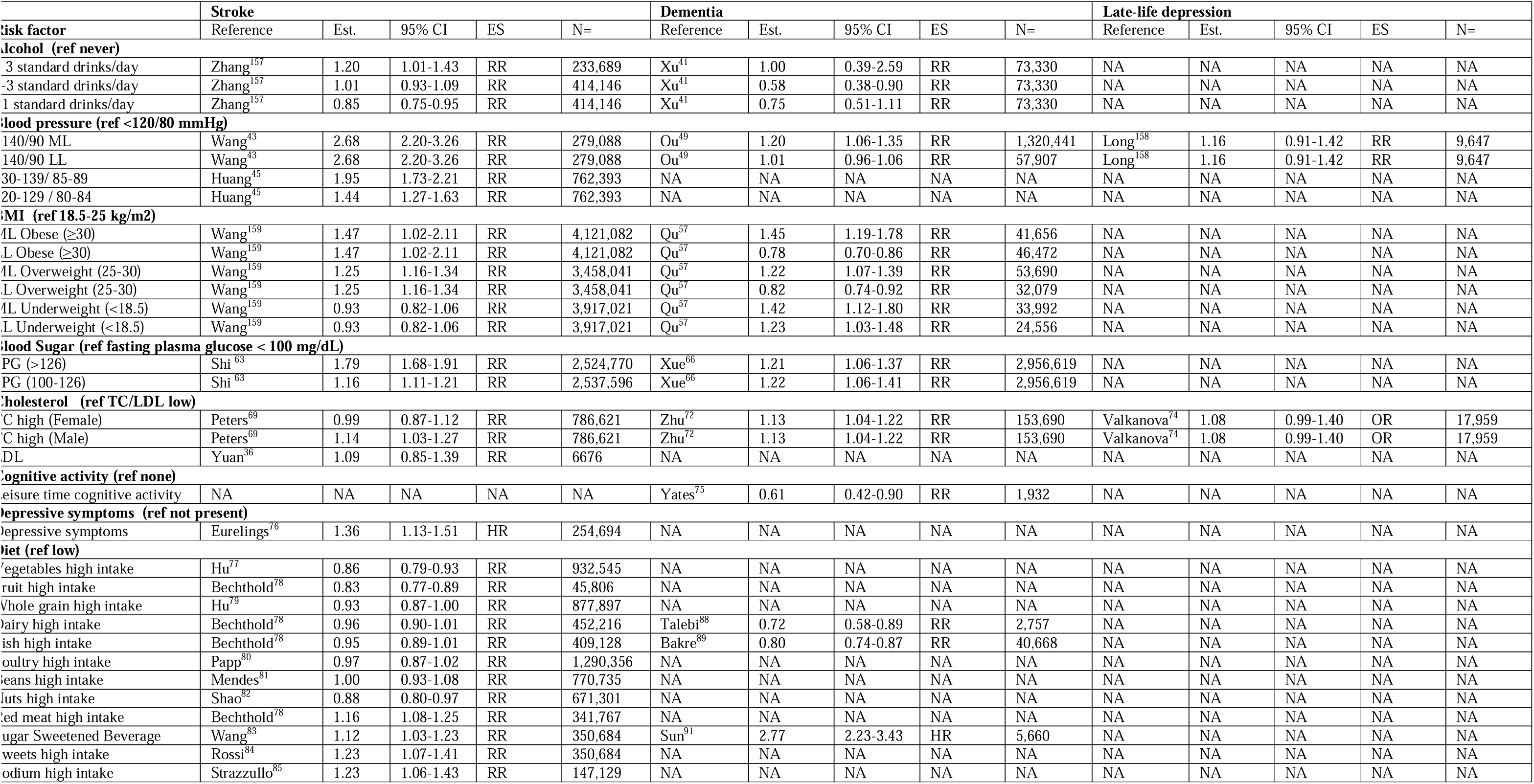
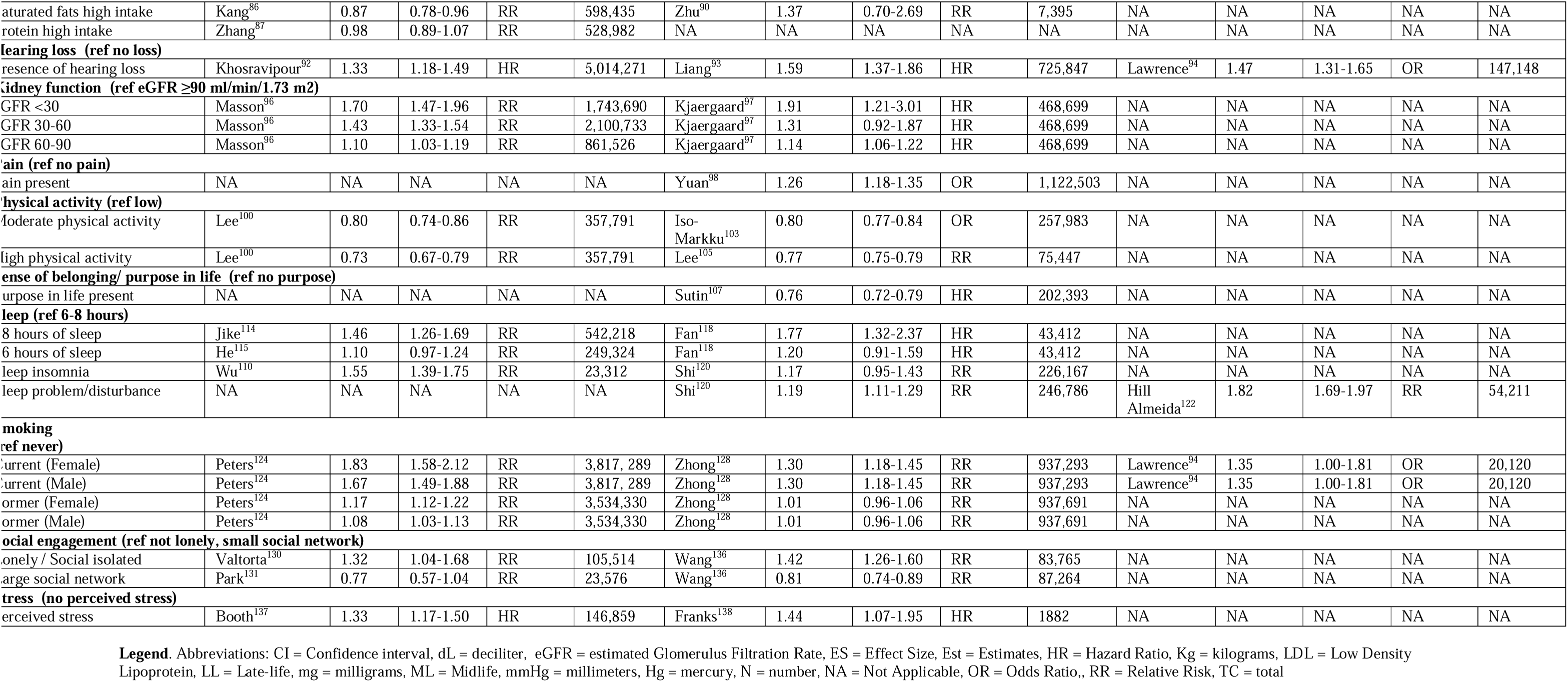
Selected studies and identification of the most relevant effect size.

### DALY weighted risk factors for a composite outcome

The calculalted DALY weighted effect sizes for each risk factors are presented in Table 2 and Figure 3 (intermediate calculations Table S7). Highest risk of age-related brain disease were found in hypertension, with a normalized beta of 87 and severe kidney disease (eGFR<30 ml/min/1.73m^2^), with a normalized beta of 60. The highest protective effect sizes were found in leisure time cognitive activity, with a normalized beta of -54 and high levels of physical activity with a normalized beta of - 34 (Figure 4).

**Figure 3:**
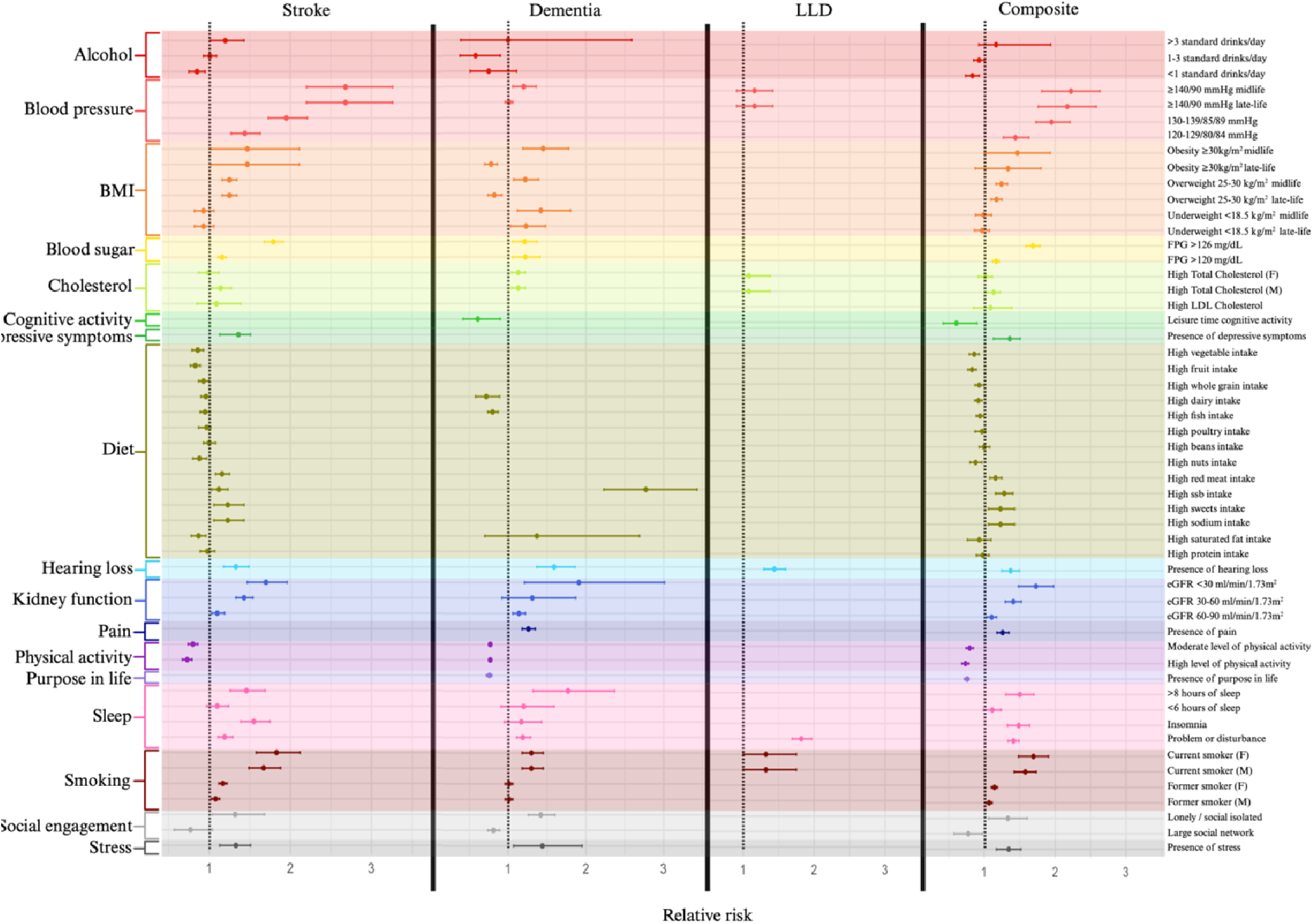
Effect sizes for stroke, dementia, LLD and a composite outcome. Abbreviations: BMI: body mass index, dL: deciliter, eGFR: estimated glomerular filtration rate, F: female, FPG: fasting plasma glucose, KG: kilograms, LLD: late-life depression, LDL: low density lipoprotein, M: male, m: meters, mg: milligram, min: minutes, mmHg: millimeters mercury. SSB: sugar sweetened beverages.

**Figure 4:**
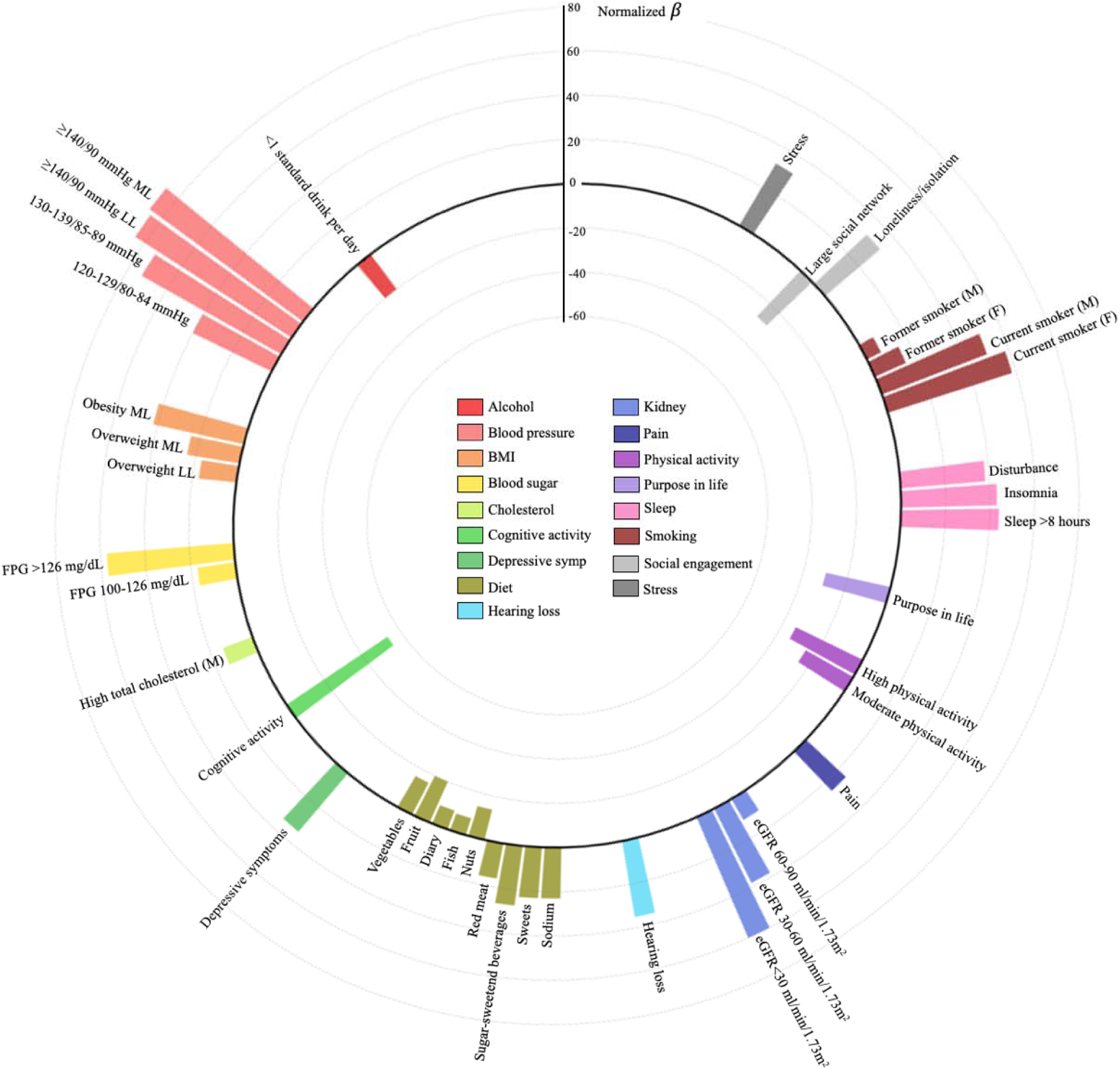
Normalized beta-coefficients for a composite outcome. Abbreviations: dL: deciliter, eGFR: estimated glomerular filtration rate, F: female, FPG: fasting plasma glucose, LL: late-life, M: male, m: meters, mg: milligram, ML: midlife min: minutes, mmHg: millimeters mercury.

**Table 2:**
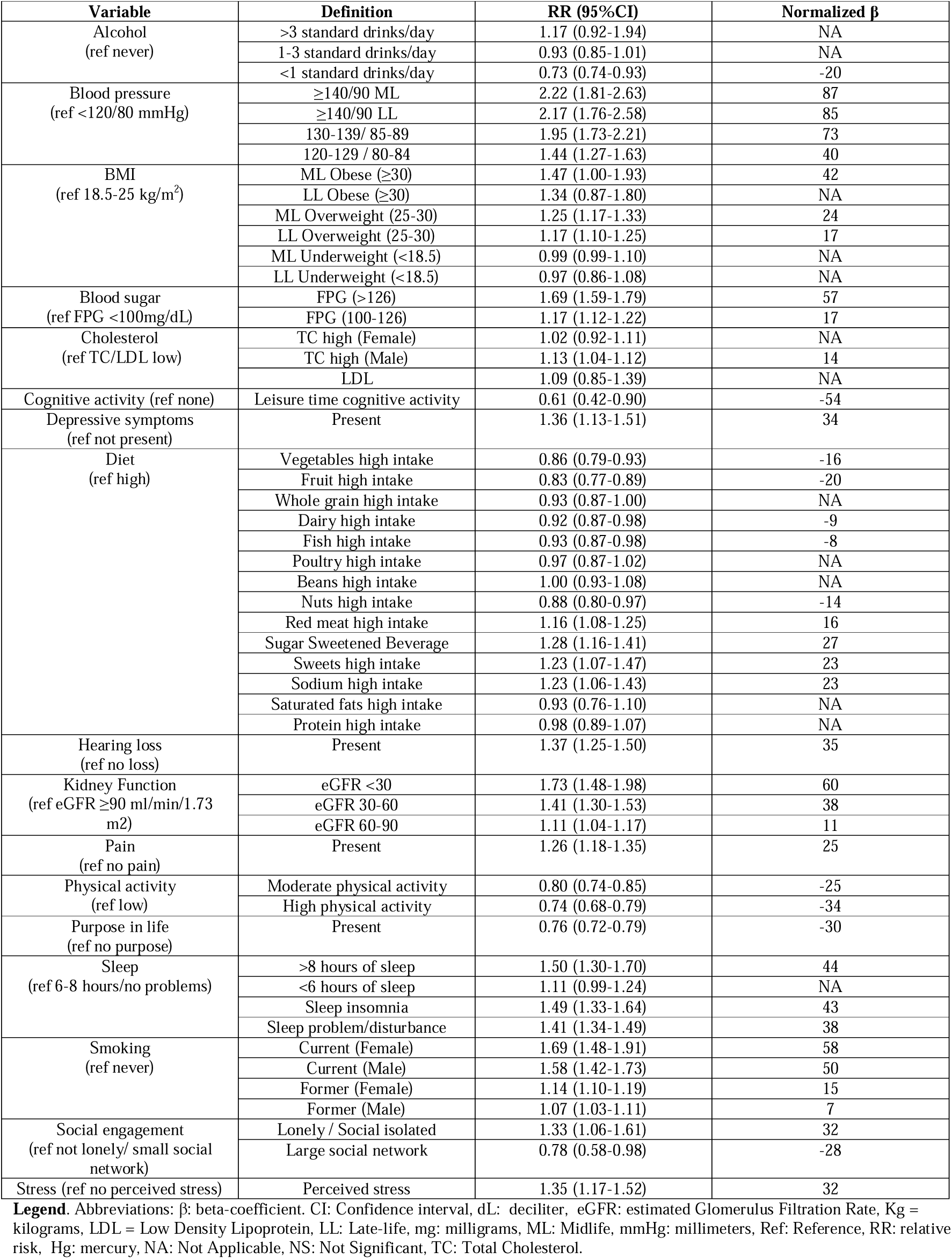
DALY weighted effect sizes for a composite outcome.

## Discussion

This study identified 18 overlapping modifiable risk factors for stroke, dementia, and LLD and calculated their relative impacts on a composite outcome using DALY-weighted beta-coefficients. By analyzing data from two systematic reviews, we weighted these risk factors according to their contribution to the burden of age-related brain diseases. This approach provided a comprehensive understanding of the relative impacts of each risk factor on the risk of a composite outcome of age-related brain disease.

When assessing individual components, hypertension was the factor with the highest individual weight. This is mainly attributable to the well-established significant association between stroke and hypertension^3,139^, and is reinforced by our methods’ DALY-based weighting system, which assigns greater impact to stroke compared to dementia or LLD^140^. Cholsterol showed limited weight with no significant LDL effect. We focused on all strokes, not distinguishing between ischemic and hemorrhagic types, which might have attenuated the effect due to the contradicting impact of cholesterol levels across stroke subtypes^141^. Additionally, excluding meta-analysis on specific treatmens or dose-responses limited available cholesterol studies. We also revealed substantial weights of leisure time cognitive activities, purpose in life, and absence of social isolation. These weights are mainly due to their association with dementia, where reverse causality may play a significant role^142^. Further, long sleep duration emerged as a major risk factor, potentially due to its relationship with possible confounders such as obesity, hypertension, and diabetes or due to reverse causality or confounding by aging. We focused on individual dietary elements as outlined by AHA Life’s Essential 8 and the DASH diet. While we did not explore the complex interplay between these dietary components^118,143,144^, there could be an overestimation of the weights of diet if combined in a future tool that builds on the results of our analysis^145^. Furthermore, dietary comparisons were often made between the highest and lowest quartiles, limiting the clinical applicability. Pain showed a significant effect on dementia risk, likely due to the direct effects of pain and the associated reduction in physical activity^98^, as well as possible reverse causality^146^. Finally, depression was included in our analysis not only as an outcome but also as a risk factor due to its bidirectional associations with vascular brain disease ^147^. Depressive symptoms showed an increased risk of stroke, which might be due to both immunological and inflammation effects^148^, as well as its association with poor health behaviors such as smoking and physical inactivity^149^. As we did not include disease-specific populations, we did not access the associations of post-stroke and post-dementia depression^150,151^.

Some limitations should be considered when interpreting our results. First, we only included overlapping risk factors across the three age-related brain diseases, thereby possibly excluding important modifiable risk factors for an individual disease, such as personality attributes and maladaptive thoughts and behaviors for LLD^152^. However, emerging evidence shows similarities in the biological pathology of dementia, stroke, and LLD, particularly due to small vessel disease - which subsequently has overlapping modifiable risk factors^3,9,153^. Second, potential interactions between risk factors, such as interactions or collinearity, were not considered^143,154^. Third, variations in definitions across different meta-analyses were encountered during our weighted risk factor calculations, which limited our use of these categorizations. Fourth, the potential for bias (including reverse causality, particularly important for dementia with its extended prodromal stages) and confounding presents a significant challenge and may have influenced the effect sizes^155^. Fifth, we limited inclusion criteria to articles written in English. Finally, the limited amount of published meta-analyses on LLD risk factors could affect our risk factor weight calculations’ overall comprehensiveness, and representativeness.

Given the limited meta-analyses on modifiable risk factors for LLD, future research should prioritize producing high-quality meta-analyses in this area. The overlap in modifiable risk factors presents an opportunity to simultaneously reduce the risk of stroke, dementia, and LLD. Developing holistic tools or models that effectively address these factors could facilitate the prevention and management of age-related brain diseases^10,24,25^. This study provided insights into the relative impact of modifiable risk factors, filling a critical gap in the literature necessary for building such comprehensive tools or models. Future research can use our findings as an empirical foundation for building such tools.

Future studies should explore certain areas that our current calculations have not fully resolved, to enhance the development of a tool or model ready for validation. These areas include assessing the utility and feasibility of the findings, which should incorporate an evaluation of social determinants of health, as well as refining the calibration process. These shortcomings could be addressed in a Delphi process^156^, ensuring the development of a comprehensive tool that leverages all available evidence to empower, educate, and motivate patients and practitioners to adopt lifestyle changes and reduce the risk of stroke, dementia, and LLD^13^.

## Conclusion

In this study, we systematically identified overlapping modifiable risk factors and calculated the relative impact of these factors on the risk of a composite outcome of stroke, dementia, and depression. These findings could guide preventative strategies and could serve as an empirical foundation for future development of tools that can empower people to change modifiable risk factors associated with these diseases.

## Data Availability

This manuscript only utilized previously published data. All data employed in this study, including the intermediate calculations, are comprehensively presented in the supplementary tables accompanying this manuscript. Should there be any inquiries or requests for further clarification regarding the data, please contact the corresponding author.

https://www.zotero.org/groups/5402286/sroma/collections/PY6XGESM

## Acknowledgment

Funding: US National Institutes of Health and American Heart Association.

