## Supplementary Material for "Modifiable Risk Factors for Stroke, Dementia, and Late-Life Depression: A Systematic Review and DALY Weighted Risk Factors for a Composite Outcome"

**Affiliations:**

**Running head**: Modifiable Risk Factors for Brain Disease

**Corresponding author:**

- Sanjula Dhillon Singh
- Address: McCance Center for Brain Health, Massachusetts General Hospital, Harvard Medical School, 399 Revolution Drive, Sommerville, MA 02145, United States of America

**Supplementary Material**

**Figures**

**Figure S1:** Prisma Flowchart of the Literature Search for risk factor identification

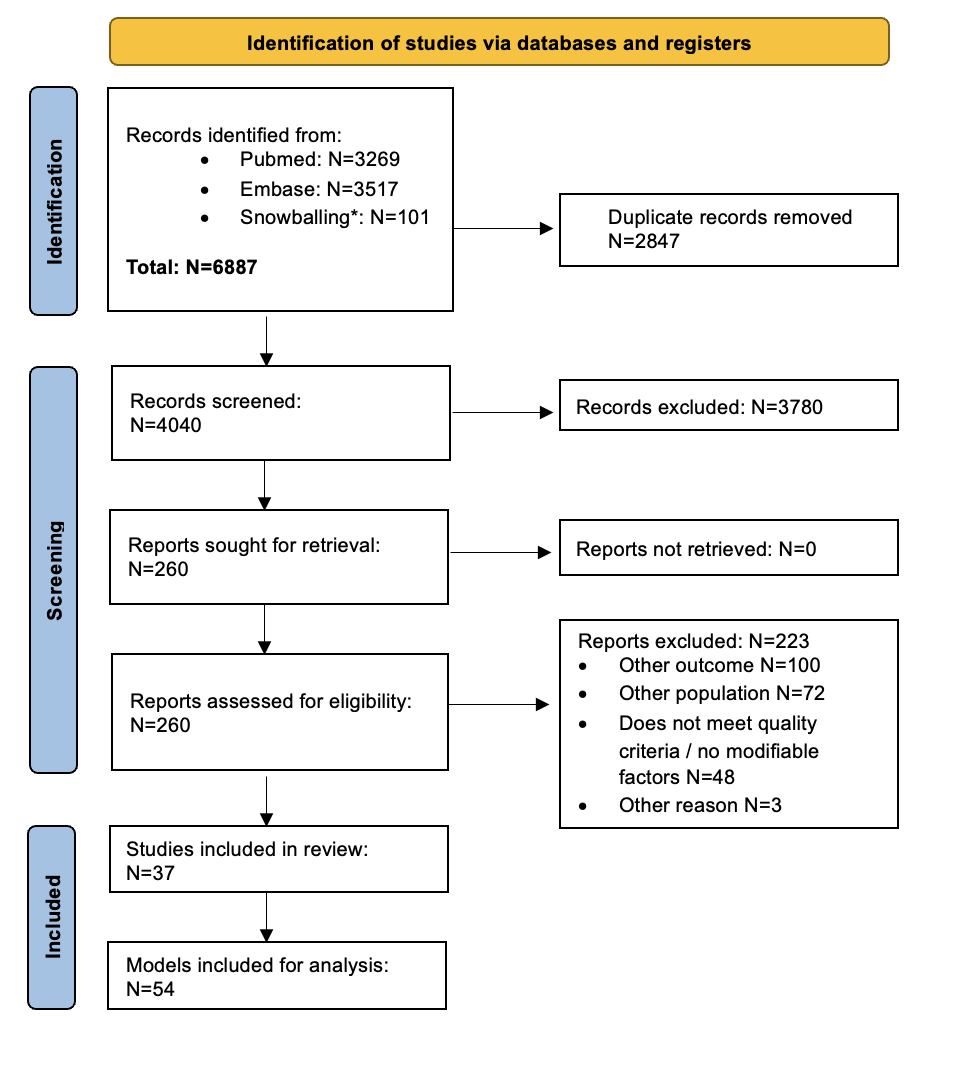

**Legend.** N denotes the number of articles. * Snowballing: new literature is identified by examining references in initially found articles, ensuring a comprehensive search.

**Tables**

**Table S1:** Search syntax literature search for risk factor identification

| **#** | **Search query Pubmed** | **Results** |
| --- | --- | --- |
| 1 | ("depressive disorder, major"[MeSH Terms] OR "Late-life depression"[All Fields] OR "late-onset depression"[All Fields] OR "late-life depressive disorder"[All Fields] OR "geriatric depression"[All Fields] OR "Depression in older adults"[All Fields] OR "Late-onset depressive symptoms"[All Fields] OR "depression in the elderly"[All Fields] OR "Late-life mood disorders"[All Fields] OR “Dementia”[MeSH Terms] OR "alzheimer disease"[MeSH Terms] OR "dementia, vascular"[MeSH Terms] OR "frontotemporal lobar degeneration"[MeSH Terms] OR "lewy body disease"[MeSH Terms] OR "Mixed Dementias"[MeSH Terms] OR "stroke"[MeSH Terms] OR "ischemic attack, transient"[MeSH Terms] OR "stroke, lacunar"[MeSH Terms] OR "cerebral hemorrhage"[MeSH Terms] OR "subarachnoid hemorrhage"[MeSH Terms] OR "cerebrovascular disorders"[MeSH Terms] OR "Brain Health"[All Fields]) | 672,331 |
| 2 | #1 AND ("risk factors"[MeSH Terms] OR "risk assessment" [MeSH Terms) | 72,983 |
| 3 | #2 AND ("Score*" [All fields] OR "Index*" [All fields] OR "Indices" [All fields] OR Model* [All fields]) | 37,007 |
| 4 | #3 AND ("practice guideline"[Publication Type] OR "review"[Publication Type] OR "systematic review"[Filter] OR "validation study"[Publication Type]) AND 2000/01/01:2023/12/31[Date - Publication]) AND (2000/1/1:2023/7/1[pdat]) | 3,269 |
| **#** | **Search query Embase** | **Results** |
| 1 | 'major depressive disorder'/exp OR 'late-life depression':ti,ab OR 'late-onset depression':ti,ab OR 'late-life depressive disorder':ti,ab OR 'geriatric depression':ti,ab OR 'depression in older adults':ti,ab OR 'late-onset depressive symptoms':ti,ab OR 'depression in the elderly':ti,ab OR 'late-life mood disorders':ti,ab OR 'dementia'/exp OR 'alzheimer disease'/exp OR 'vascular dementia'/exp OR 'frontotemporal dementia'/exp OR 'dementia with lewy bodies'/exp OR 'mixed dementias':ti,ab OR 'stroke'/exp OR 'transient ischemic attack'/exp OR 'lacunar infarction'/exp OR 'cerebral hemorrhage'/exp OR 'subarachnoid hemorrhage'/exp OR 'cerebrovascular disorders'/exp OR 'brain health':ti,ab | 1,399,065 |
| 2 | 'risk factor'/exp OR 'risk assessment'/exp | 1,899,937 |
| 3 | score*:ti,ab OR index*:ti,ab OR indices:ti,ab OR model*:ti,ab | 7,440,016 |
| 4 | #1 AND #2 AND #3 | 63,317 |
|  | #4 AND [01-01-2000]/sd NOT [02-07-2023]/sd AND ('practice guideline'/de OR 'systematic review'/de OR 'validation process'/de OR 'validation study'/de) AND ('article'/it OR 'review'/it) | 3517 |

**Table S2:** Search syntax systematic review of meta-analyses.

Pubmed

| **#** | **Search query Pubmed** | **Results** |
| --- | --- | --- |
| 1 | ("Blood pressure"[MeSH Terms] OR "Blood pressure"[All Fields] OR "Systolic blood pressure"[All Fields] OR "diastolic blood pressure"[All Fields] OR "Pulse Pressure"[All Fields] OR "Arterial Pressure"[MeSH Terms] OR "Hypertension"[MeSH Terms] OR "high blood pressure*"[All Fields] OR ("Body mass index"[MeSH Terms] OR "Body mass index"[All Fields] OR "Body Weight"[MeSH Terms] OR "Overweight"[MeSH Terms] OR "Obesity"[MeSH Terms] OR "Thinness"[MeSH Terms] OR "Overweight"[All Fields] OR "Adiposity"[MeSH Terms] OR "Adiposity"[All Fields] OR "Underweight"[All Fields]) OR (((("Cholesterol"[MeSH Terms] OR "Lipids"[MeSH Terms] OR "Lipoproteins"[MeSH Terms] OR "LDL"[All Fields] OR "HDL"[All Fields] OR "Total cholesterol"[All Fields]) AND "Hyperlipidemias"[MeSH Terms]) AND "lipemia*"[All Fields]) OR "lipidemia*"[All Fields] OR "hyperchol*"[All Fields] OR "hyperlipo*"[All Fields] OR "hypertrigly*"[All Fields] OR "dyslipi*"[All Fields]) OR ("Diabetes Mellitus"[MeSH Terms] OR "diabet*"[All Fields] OR "Glucose intolerance"[MeSH Terms] OR "Hyperglycemia"[MeSH Terms] OR "Impaired Glucose Tolerance"[All Fields] OR "glucose intolerance*"[All Fields] OR "Diabetes Mellitus Type 1"[All Fields] OR "Diabetes Mellitus Type 2"[All Fields] OR "glycated hemoglobin"[MeSH Terms] OR "HbA1c"[All Fields] OR "Glycated Hemoglobin A"[All Fields]) OR ("Renal Insufficiency"[MeSH Terms] OR "renal insufficien*"[All Fields] OR "kidney insufficien*"[All Fields] OR "insufficiency kidney"[All Fields] OR "kidney fail*"[All Fields] OR "renal fail*"[All Fields]) OR ("Hearing Loss"[MeSH Terms] OR "loss hearing"[All Fields] OR "hypoacus*"[All Fields] OR "hearing impair*"[All Fields] OR "transitory deaf*"[All Fields] OR "transitory hearing loss*"[All Fields] OR "Presbycusis"[MeSH Terms] OR "Presbycusis"[All Fields]) OR ("depressive symptom*"[All Fields] OR "symptom depressive"[All Fields] OR "emotional depress*"[All Fields] OR "Affective Symptoms"[MeSH Terms] OR "Mood disturbance"[All Fields] OR "feeling blue*"[All Fields] OR "feeling down*"[All Fields]) OR ("Pain"[MeSH Terms] OR "burning pain*"[All Fields] OR "physical suffer*"[All Fields] OR "migratory pain*"[All Fields] OR "radiating pain*"[All Fields] OR "splitting pain*"[All Fields] OR "ache*"[All Fields] OR "crushing pain*"[All Fields]) OR ("Smoking"[MeSH Terms] OR "smoking behavior*"[All Fields] OR "smoking habit*"[All Fields] OR "Tobacco Use"[MeSH Terms] OR "tobacco use*"[All Fields] OR "tobacco consumption*"[All Fields] OR "tobacco chewing*"[All Fields] OR "Cigarette Smoking"[MeSH Terms]) OR ("Alcohol Drinking"[MeSH Terms] OR "drinking alcohol*"[All Fields] OR "alcohol consumption*"[All Fields] OR "alcohol intake*"[All Fields] OR "alcohol drinking habit*"[All Fields] OR "Alcoholism"[MeSH Terms] OR "alcohol dependence*"[All Fields] OR "alcohol addiction*"[All Fields] OR "alcoholic intoxication chronic*"[All Fields] OR "alcohol abuse*"[All Fields] OR "ethanol abuse*"[All Fields] OR "alcohol use disorder*"[All Fields]) OR ("Exercise"[MeSH Terms] OR "exercis*"[All Fields] OR "phys activity"[All Fields] OR "physical exercis*"[All Fields] OR "acute exercise*"[All Fields] OR "isometric exercise*"[All Fields] OR "aerobic exercise"[All Fields] OR "exercise training*"[All Fields] OR "sport*"[All Fields] OR "athletic*"[All Fields]) OR ((((("Sleep"[MeSH Terms] OR "Sleep Quality"[MeSH Terms] OR "sleep habit"[All Fields] OR "Sleep Wake Disorder"[All Fields]) AND "sleep wake disorder*"[All Fields]) OR "sleep disorder*"[All Fields] OR "short sleeper syndrome*"[All Fields] OR "short sleep phenotype*"[All Fields]) AND "sleep disorders, intrinsic"[MeSH Terms]) OR "intrinsic sleep disorder*"[All Fields] OR "sleep state misperception*"[All Fields] OR "hypersomnia*"[All Fields] OR "Dyssomnias"[MeSH Terms] OR "dyssomnia*"[All Fields] OR "extrinsic sleep disorder*"[All Fields] OR "limit setting sleep disorder*"[All Fields] OR "nocturnal eating drinking syndrome*"[All Fields] OR "adjustment sleep disorder*"[All Fields] OR "environmental sleep disorder*"[All Fields]) OR ("stress, psychological"[MeSH Terms] OR "Life stress"[All Fields] OR "Psychological stressors"[All Fields] OR "financial stress*"[All Fields] OR "financial pressure"[All Fields] OR "financial toxicity"[All Fields] OR "economic burden*"[All Fields] OR "job stress*"[All Fields] OR "work related stress*"[All Fields] OR "occupational stress*"[MeSH Terms]) OR ((("social support"[MeSH Terms] OR "Social Participation"[MeSH Terms]) AND "social support"[All Fields]) OR "Social Isolation"[All Fields] OR "social exclusion"[All Fields] OR "Social integration"[All Fields] OR "Social interaction"[All Fields] OR "Interpersonal relations"[All Fields] OR "participation social"[All Fields] OR "social engage*"[All Fields] OR "engagement social"[All Fields] OR "Social Citizenship"[All Fields] OR "citizenship social"[All Fields]) OR ((("self rated health"[All Fields] OR "self rated health"[All Fields]) AND "concept self"[All Fields]) OR "self perception*"[All Fields] OR "self perception*"[All Fields]) OR ("Purpose in life"[All Fields] OR "Meaning in life"[All Fields] OR "Sense of belonging"[All Fields] OR "Satisfaction in Life"[All Fields]) OR ("Cognitive Training"[MeSH Terms] OR "training cognitive"[All Fields] OR "cognitive train*"[All Fields] OR "brain train*"[All Fields] OR "memory train*"[All Fields] OR "cognitive activit*"[All Fields] OR "puzzl*"[All Fields]) OR ("diet*"[MeSH Terms] OR "vegetable*"[All Fields] OR "vegetable intake"[All Fields] OR "fruit intake"[All Fields] OR "fruit*"[All Fields] OR "diet, healthy"[MeSH Terms] OR "healthy diet*"[All Fields] OR "healthy eating*"[All Fields] OR "healthy nutrition*"[All Fields] OR "prudent diet*"[All Fields] OR "healthy eating index*"[All Fields] OR "fish*"[All Fields] OR "fish intake"[All Fields] OR "diet, mediterranean"[MeSH Terms] OR "nutritional requirement*"[All Fields] OR "nutrition requirement*"[All Fields] OR "dietary requirement*"[All Fields])) | 6,826,548 |
| 2 | "Late-life depression"[All Fields] OR "late-onset depression"[All Fields] OR "late-life depressive disorder"[All Fields] OR "geriatric depression"[All Fields] OR "Depression in older adults"[All Fields] OR "Late-onset depressive symptoms"[All Fields] OR "depression in the elderly"[All Fields] OR "Late-life mood disorders"[All Fields] OR "Dementia"[MeSH Terms] OR "alzheimer disease"[MeSH Terms] OR "dementia, vascular"[MeSH Terms] OR "frontotemporal lobar degeneration"[MeSH Terms] OR "Mixed Dementias"[MeSH Terms] OR "stroke"[MeSH Terms] OR "ischemic attack, transient"[MeSH Terms] OR "stroke, lacunar"[MeSH Terms] OR "cerebral hemorrhage"[MeSH Terms] OR "cerebrovascular disorders"[MeSH Terms] | 632,115 |
| 3 | "meta analysis" [Title] OR "meta-analysis" [Title] OR "meta analyses" [Title] OR "meta-analyses" [Title] | 185,059 |
| 4 | #1 AND #2 AND #3  AND ((humans[Filter]) AND (2000:2023[pdat])) | 2,084 |

Embase

| **#** | **Search query Embase** | **Results** |
| --- | --- | --- |
| 1 | 'blood pressure'/exp/mj OR 'blood pressure':ti,ab OR 'systolic blood pressure':ti,ab OR 'diastolic blood pressure':ti,ab OR 'pulse pressure':ti,ab OR 'arterial pressure'/exp/mj OR 'hypertension'/exp/mj OR 'high blood pressure*':ti,ab OR 'body mass'/exp/mj OR 'body mass index':ti,ab OR 'body weight'/exp/mj OR 'overweight':ti,ab OR 'obesity'/exp/mj OR 'underweight'/exp/mj OR 'thinness':ti,ab OR 'adiposity':ti,ab OR 'cholesterol'/exp/mj OR 'lipid'/exp/mj OR 'lipid*':ti,ab OR 'lipoprotein'/exp/mj OR 'low density lipoprotein'/exp/mj OR 'low density lipoprotein cholesterol'/exp/mj OR 'high density lipoprotein'/exp/mj OR 'high density lipoprotien' OR 'cholesterol blood level'/exp/mj OR 'hyperlipidemia'/exp/mj OR 'lipemia':ti,ab OR 'lipidemia':ti,ab OR 'dyslipidemia'/exp/mj OR 'hypercholesterolemia'/exp/mj OR 'hypertriglyceridemia'/exp/mj OR 'diabetes mellitus'/exp/mj OR 'diabetes':ti,ab OR 'glucose intolerance'/exp/mj OR 'hyperglycemia'/exp/mj OR 'impaired glucose tolerance'/exp/mj OR 'insulin dependent diabetes mellitus'/exp/mj OR 'non insulin dependent diabetes mellitus'/exp/mj OR 'hemoglobin a1c'/exp/mj OR 'glycated hemoglobin'/exp/mj OR 'kidney failure'/exp/mj OR 'chronic kidney failure'/exp/mj OR 'renal insufficien*' OR 'hearing impairment'/exp/mj OR 'hearing loss':ti,ab OR 'perception deafness'/exp/mj OR 'conduction deafness'/exp/mj OR 'hypoacusia'/exp/mj OR 'presbyacusis'/exp/mj OR 'depressive symptoms':ti,ab OR 'depression'/exp/mj OR 'emotional disorder'/exp/mj OR 'mood disorder' OR 'feeling blue':ti,ab OR 'feeling down':ti,ab OR 'cognitive rehabilitation'/exp/mj OR 'cognitive training':ti,ab OR 'brain training':ti,ab OR 'memory training'/exp/mj OR 'cognitive activity' OR 'puzzles' OR 'pain'/exp/mj OR 'physical suffering' OR 'smoking'/exp/mj OR 'smoking behaviour':ti,ab OR 'smoking habit'/exp/mj OR 'tobacco use'/exp/mj OR 'tobacco dependence'/exp/mj OR 'cigarette smoking'/exp/mj OR 'alcohol'/exp/mj OR 'alcohol drinking'/exp/mj OR 'alcohol habit':ti,ab OR 'alcoholism'/exp/mj OR 'alcohol intoxication'/exp/mj OR 'ethanol'/exp/mj OR 'ethanol abuse':ti,ab OR 'alcohol abuse'/exp/mj OR 'exercise'/exp/mj OR 'physical activity'/exp/mj OR 'isometric exercise'/exp/mj OR 'aerobic exercise' OR 'sport'/exp/mj OR 'athletics'/exp/mj OR 'diet'/exp/mj OR 'nutrition'/exp/mj OR 'dietary intake'/exp/mj OR 'vegetable*' OR 'vegetable consumption'/exp/mj OR 'fruit'/exp/mj OR 'fruit consumption'/exp/mj OR 'healthy diet'/exp/mj OR 'healthy diet score'/exp/mj OR 'healthy eating index'/exp/mj OR 'fish*' OR 'fish consumption'/exp/mj OR 'mediterranean diet'/exp/mj OR 'nutritional requirement'/exp/mj OR 'dietary requirement':ti,ab OR 'sleep'/exp/mj OR 'sleep disorder'/exp/mj OR 'sleep disturbance':ti,ab OR 'sleep quality'/exp/mj OR 'sleep wake disorder':ti,ab OR 'short sleeper syndrom':ti,ab OR 'intrinsic sleep disorder'/exp/mj OR 'hypersomina' OR 'extrinsic sleep disorder':ti,ab OR 'dyssomnia'/exp/mj OR 'mental stress'/exp/mj OR 'psychological stress'/exp/mj OR 'life stress'/exp/mj OR 'financial stress'/exp/mj OR 'job stress'/exp/mj OR 'occupational stress':ti,an OR 'social support'/exp/mj OR 'social deprivation'/exp/mj OR 'social alienation'/exp/mj OR 'solitary confinement'/exp/mj OR 'loneliness'/exp/mj OR 'social interaction'/exp/mj OR 'integration'/exp/mj OR 'social engagement'/exp/mj OR 'social citizenship' OR 'self rated health'/exp/mj OR 'self concept'/exp/mj OR 'self perception':ti,ab OR 'purpose in life'/exp/mj OR 'meaning in life'/exp/mj OR 'sense of belonging'/exp/mj OR 'satisfaction in life' | 7,077,853 |
| 2 | 'geriatric depression'/exp/mj OR 'late onset depression'/exp/mj OR 'late-life depression'/exp/mj OR 'alzheimer* disease' OR 'frontotemporal dementia'/exp/mj OR 'mixed dementia'/exp/mj OR 'multiinfarct dementia'/exp/mj OR 'cortical sclerosis, diffuse'/exp/mj OR 'alzheimer neuron degeneration'/exp/mj OR 'alzheimer, dementia'/exp/mj OR 'frontal dementia'/exp/mj OR 'frontal lobe dementia'/exp/mj OR 'mixed dementia*' OR 'vascular dementia'/exp/mj OR 'cerebrovascular accident'/exp/mj OR 'brain hemorrhage'/exp/mj OR 'brain hematoma'/exp/mj OR 'brain ischemia'/exp/mj OR 'transient ischemic attack'/exp/mj OR 'stroke'/exp/mj OR 'cva'/exp/mj OR 'intracerebral hemorrhage'/exp/mj OR 'intracerebral ischemic attack' | 604,324 |
| 3 | 'meta analysis':ti OR 'meta-analysis':ti OR 'meta-analyses':ti OR 'meta analyses':ti | 223,123 |
| 4 | #1 AND #2 AND #3  AND [embase]/lim AND 'human'/de AND ('article'/it OR 'review'/it) | 1,136 |

PsycInfo

| **#** | **Search query Embase** | **Results** |
| --- | --- | --- |
| 1 | ((Blood pressure) OR (Systolic blood pressure) OR (diastolic blood pressure) OR (Pulse Pressure) OR (Arterial Pressure) OR (Hypertension) OR (high blood pressure*) OR (Body mass index) OR (Body Weight) OR (Overweight) OR (Obesity) OR (Thinness) OR (Adiposity) OR (Underweight) OR (Cholesterol) OR (Lipids) OR (Lipoproteins) OR (LDL) OR (HDL) OR (Total cholesterol) OR (Hyperlipidemias) OR (lipemia*) OR (lipidemia*) OR (hyperchol*) OR (hyperlipo*) OR (hypertrigly*) OR (dyslipi*) OR (Diabetes Mellitus) OR (diabet*) OR (Glucose intolerance) OR (Hyperglycemia) OR (Impaired Glucose Tolerance) OR (glucose intolerance*) OR (Diabetes Mellitus Type 1) OR (Diabetes Mellitus Type 2) OR (glycated hemoglobin) OR (HbA1c) OR (Glycated Hemoglobin A) OR (Renal Insufficiency) OR (renal insufficien*) OR (kidney insufficien*) OR (insufficiency kidney) OR (kidney fail*) OR (renal fail*) OR (Hearing Loss) OR (loss hearing) OR (hypoacus*) OR (hearing impair*) OR (transitory deaf*) OR (transitory hearing loss*) OR (Presbycusis) OR (depressive symptom*) OR (symptom depressive) OR (emotional depress*) OR (Affective Symptoms) OR (Mood disturbance) OR (feeling blue*) OR (feeling down*) OR (Pain) OR (burning pain*) OR (physical suffer*) OR (migratory pain*) OR (radiating pain*) OR (splitting pain*) OR (ache*) OR (crushing pain*) OR (Smoking) OR (smoking behavior*) OR (smoking habit*) OR (Tobacco Use) OR (tobacco use*) OR (tobacco consumption*) OR (tobacco chewing*) OR (Cigarette Smoking) OR (Alcohol Drinking) OR (drinking alcohol*) OR (alcohol consumption*) OR (alcohol intake*) OR (alcohol drinking habit*) OR (Alcoholism) OR (alcohol dependence*) OR (alcohol addiction*) OR (alcoholic intoxication chronic*) OR (alcohol abuse*) OR (ethanol abuse*) OR (alcohol use disorder*) OR (Exercise) OR (exercis*) OR (phys activity) OR (physical exercis*) OR (acute exercise*) OR (isometric exercise*) OR (aerobic exercise) OR (exercise training*) OR (sport*) OR (athletic*) OR (Sleep) OR (Sleep Quality) OR (sleep habit) OR (Sleep Wake Disorder) OR (sleep wake disorder*) OR (sleep disorder*) OR (short sleeper syndrome*) OR (short sleep phenotype*) OR (sleep disorders, intrinsic) OR (intrinsic sleep disorder*) OR (sleep state misperception*) OR (hypersomnia*) OR (Dyssomnias) OR (dyssomnia*) OR (extrinsic sleep disorder*) OR (limit setting sleep disorder*) OR (nocturnal eating drinking syndrome*) OR (adjustment sleep disorder*) OR (environmental sleep disorder*) OR (stress, psychological) OR (Life stress) OR (Psychological stressors) OR (financial stress*) OR (financial pressure) OR (financial toxicity) OR (economic burden*) OR (job stress*) OR (work related stress*) OR (occupational stress*) OR (social support) OR (Social Participation) OR (Social Isolation) OR (social exclusion) OR (Social integration) OR (Social interaction) OR (Interpersonal relations) OR (participation social) OR (social engage*) OR (engagement social) OR (Social Citizenship) OR (citizenship social) OR (self rated health) OR (concept self) OR (self perception*) OR (Purpose in life) OR (Meaning in life) OR (Sense of belonging) OR (Satisfaction in Life) OR (Cognitive Training) OR (training cognitive) OR (cognitive train*) OR (brain train*) OR (memory train*) OR (cognitive activit*) OR (puzzl*) OR (diet*) OR (vegetable*) OR (vegetable intake) OR (fruit intake) OR (fruit*) OR (diet, healthy) OR (healthy diet*) OR (healthy eating*) OR (healthy nutrition*) OR (prudent diet*) OR (healthy eating index*) OR (fish*) OR (fish intake) OR (diet, mediterranean) OR (nutritional requirement*) OR (nutrition requirement*) OR (dietary requirement*))  ((Late-life depression) OR (late-onset depression) OR (late-life depressive disorder) OR (geriatric depression) OR (Depression in older adults) OR (Late-onset depressive symptoms) OR (depression in the elderly) OR (Late-life mood disorders) OR (Dementia) OR (alzheimer disease) OR (dementia, vascular) OR (frontotemporal lobar degeneration) OR (Mixed Dementias) OR (stroke) OR (ischemic attack, transient) OR (stroke, lacunar) OR (cerebral hemorrhage) OR (cerebrovascular disorders))  (title: meta-analysis)  #1 AND #2 AND #3 | 84,013 |
| 2 | 'geriatric depression'/exp/mj OR 'late onset depression'/exp/mj OR 'late-life depression'/exp/mj OR 'alzheimer* disease' OR 'frontotemporal dementia'/exp/mj OR 'mixed dementia'/exp/mj OR 'multiinfarct dementia'/exp/mj OR 'cortical sclerosis, diffuse'/exp/mj OR 'alzheimer neuron degeneration'/exp/mj OR 'alzheimer, dementia'/exp/mj OR 'frontal dementia'/exp/mj OR 'frontal lobe dementia'/exp/mj OR 'mixed dementia*' OR 'vascular dementia'/exp/mj OR 'cerebrovascular accident'/exp/mj OR 'brain hemorrhage'/exp/mj OR 'brain hematoma'/exp/mj OR 'brain ischemia'/exp/mj OR 'transient ischemic attack'/exp/mj OR 'stroke'/exp/mj OR 'cva'/exp/mj OR 'intracerebral hemorrhage'/exp/mj OR 'intracerebral ischemic attack' | 4660 |
| 3 | 'meta analysis':ti OR 'meta-analysis':ti OR 'meta-analyses':ti OR 'meta analyses':ti | 1534 |
| 4 | #1 AND #2 AND #3  AND [embase]/lim AND 'human'/de AND ('article'/it OR 'review'/it) | 38 |

**Table S3.** Overview of the formulas

| **#** | **Formula** | **Explanation** |
| --- | --- | --- |
| 1 | $RR=\frac{1-\mathcal{e}^{HR \times Ln(1-r)}}{r}$  $Lower CI RR=\frac{1-\mathcal{e}^{\log(Lower CI of HR) x Ln(1-r)}}{r}$  $Upper CI RR=\frac{1-\mathcal{e}^{\log(Upper CI of HR) x Ln(1-r)}}{r}$ | Hazard Ratio to Relative Risk^35^ |
| 2 | $RR= \frac{OR}{\left( 1-r \right)+(r \times OR)}$  $Lower CI of RR= \frac{log(Lower CI of OR)}{\left( 1-r \right)+(r \times\log(Lower CI of OR))}$  $Upper CI of RR= \frac{log(Upper CI of OR)}{\left( 1-r \right)+(r \times\log(Lower CI of OR))}$ | Odds Ratio to Relative Risk^35^ |
| 3 | ${RR}_{combined}=\sum_{i=1}^{n} {RR}_{i}\times\frac{{DALY}_{i}}{Total DALYs for all included outcomes}$ | Combined Relative Risk^2^ |
| 6 | $Var\left( {RR}_{i} \right)=\left( \frac{{CI Upper}_{i}- {CI lower}_{i}}{3.92} \right)^{2}$  $Var({RR}_{combined})=\sum_{i=1}^{n} {(Var}_{i}\times({\frac{{DALY}_{i}}{Total DALYs for all included outcomes})}^{2}$  SE$\left( {RR}_{combined} \right)= \sqrt{Var(({RR}_{combined})}$  $Lower CI\left( {RR}_{combined} \right)={RR}_{combined}-1.96* \mathrm{SE}\left( {RR}_{combined} \right)$  $Upper CI\left( {RR}_{combined} \right)={RR}_{combined}+1.96* \mathrm{SE}\left( {RR}_{combined} \right)$ | Combined 95% Confidence interval^159^ |
| 5 | *𝛽=*Ln*(RR)* | Relative Risk to Beta-Coefficient^15^ |
| 6 | $scaled \beta_{i} =\beta_{i} \times\frac{1}{Lowest \beta}$ | Beta-Coefficient to Scaled Beta-Coefficient^15^ |

Abbreviations: β = Beta-coefficient, CI = Confidence Interval, Cm = Care model, DALY = Disability adjusted life years, e = Euler's number, HR = Hazard Ratio, I = index, Ln = Natural logarithm, Log = Logarithm, n = Count, OR = Odds Ratio, r = incidence rate, RR = Relative Risk, Var = Variance, Σ = Summation

**Table S4:** Overview of included models for risk factor identification

| **Model** | **First Author** | **Year of publication** | **Cohort** | **Country of cohort** | **Sample size, N=** | **Model used** | **Included risk factors** |
| --- | --- | --- | --- | --- | --- | --- | --- |
| **STROKE** | | | | | | | |
| **PROCAM**^160^ | Assman | 2007 | Prospective Cardiovascular  Münster (PROCAM) study | Germany | 45435 | Cox Proportional Hazard Model | Age, DM, SBP, Sex, Smoking. |
| **MORGAM Female**^161^ | Borglykke | 2010 | Monica Risk, Genetics, Archiving and Monograph (MORGAM) Collaboration | Nine European Countries | NA | Cox Proportional Hazard Model | BMI, DBP, DM, HDL, anti-hypertensive medication use, SBP, Smoking, Total Cholesterol. |
| **MORGAM Male**^161^ | Borglykke | 2010 | Monica Risk, Genetics, Archiving and Monograph (MORGAM) Collaboration | Nine European Countries | NA | Cox Proportional Hazard Model | BMI, DBP, DM, HDL, anti-hypertensive medication use, SBP, Smoking, Total Cholesterol. |
| **Qbleed Female**^162^ | Chambless | 2004 | Atherosclerosis Risk in Communities Study (ARIC) | USA | 15792 | Cox Proportional Hazard Model | AF, Alcohol, Anti-depressant use, Anticoagulant use, Antiplatelet use, Carbamazepine use, Chronic liver disease, Ethnicity, Esophagus varices, Pancreatitis, Phenytoin use, Platelet count, Previous bleeds, Sex, Smoking, Treated hypertension, Townsend score. |
| **Qbleed Male**^162^ | Chambless | 2004 | Atherosclerosis Risk in Communities Study (ARIC) | USA | 15792 | Cox Proportional Hazard Model | AF, Alcohol, Anti-depressant use, Anticoagulant use, Antiplatelet use, Carbamazepine use, Chronic liver disease, Ethnicity, Esophagus varices, Pancreatitis, Phenytoin use, Platelet count, Previous bleeds, Sex, Smoking, Treated hypertension, Townsend score. |
| **Chien, et al**^163^ | Chien | 2010 | ChinShan Community Cohort Study Taiwain | Taiwan | NA | Cox Proportional Hazard Model | AF, Age, DBP, DM, FH: stroke, Gender, SBP. |
| **Framingham Stroke Risk Score Female**^16^ | D’Agostino | 1994 | Framingham Heart Study | USA | 5734 | Cox Proportional Hazard Model | AF, Age Cardiovascular disease, DM, LVH, Sex, SBP, Smoking. |
| **Framingham Stroke Risk Score MALE**^16^ | D’Agostino | 1994 | Framingham Heart Study | USA | 5734 | Cox Proportional Hazard Model | AF, Age Cardiovascular disease, DM, LVH, Sex, SBP, Smoking. |
| **Ross**^164^ | El-Haijj | 2019 | Score Generation Study | Libya | 732 | Cox Proportional Hazard Model | AF, Age, anticoagulant use, anti-hypertensive medication use, Blood Pressure, CHD, DVT, Hypertension, Migraine, PE, Smoking. |
| **Ferket, et al. Extendend Hemorrhagic Model**^165^ | Ferket | 2014 | (i)Atherosclerosis Risk in Communities Study (ARIC)  (ii) Rotterdam Study  (iii) Cardiovascular Health Study | (i) + (iii) USA (ii) NL | (i): 15170 (ii): 6910 (iii): 5431 | Cox Proportional Hazard Model | Age, Sex, Ethnicity, Smoking, DM, Anti-hypertensive use, SBP, CHD, DBP, HDL-Total Cholesterol Ratio, Kidney Function, Waist-to-hip ratio |
| **Ferket, et al. Extendend Ischemic Model**^165^ | Ferket | 2014 | (i)Atherosclerosis Risk in Communities Study (ARIC)  (ii)Rotterdam Study  (iii)Cardiovascular Health Study | (i) + (iii) USA (ii) NL | (i): 15170 (ii): 6910 (iii): 5431 | Cox Proportional Hazard Model | Age, Anti-hypertensive use, CHD, DBP, DM, Ethnicity, HDL-Total Cholesterol Ratio, Kidney Function, SBP, Sex, Smoking, Waist-to-hip ratio. |
| **Friedland, et al Audiogram Model**^166^ | Friedland | 2009 | Medical College of Wisconsin and Froedtert Hospital | USA | 1168 | Cox Proportional Hazard Model | Age, Hearing, Hyperlipidemia, Hypertension, Smoking. |
| **Qstroke Female**^18^ | Hippisley-Cox | 2013 | National Qresearch database | UK | 3.5 million | Cox Proportional Hazard Model | AF, Anti-hypertensive Treatment, Age, BMI, CHD, CHF, CKD, deprivation score, ethnicity, FH: CHD, HDL, RA, SBP, smoking, Total Cholesterol, Type 1 DM, Type 2 DM, Valvular Heart Disease. |
| **Qstroke Male**^18^ | Hippisley-Cox | 2013 | National Qresearch database | UK | 3.5 million | Cox Proportional Hazard Model | AF, Anti-hypertensive Treatment, Age, BMI, CHD, CHF, CKD, deprivation score, ethnicity, FH: CHD, HDL, RA, SBP, smoking, Total Cholesterol, Type 1 DM, Type 2 DM, Valvular Heart Disease. |
| **SRSRF**^167^ | Howard | 2017 | REGARDS | USA | 30239 | Logistic Regression Model | AF, Age, DM, Education, Ethnicity, General self-reported health, MI, Self-reported AF, Self-reported DM, Self-reported hypertension, Sex, Smoking. |
| **Lee, et al. Female Model**^168^ | Lee | 2018 | NHS Korea | Korea | 2532986 | Cox Proportional Hazard Model | Age, Alcohol, BMI, DM, Diastolic blood pressure, Family history of heart disease, Family history of stroke, History of heart disease, Hypertension, Physical activity, Sex, Smoking, Systolic blood pressure, Total Cholesterol. |
| **Lee, et al. Male Model**^168^ | Lee | 2018 | NHS Korea | Korea | 3182325 | Cox Proportional Hazard Model | Age, Alcohol, BMI, DM, Diastolic blood pressure, Family history of heart disease, History of heart disease, Hypertension, Physical activity, Sex, Smoking, Systolic blood pressure, Total Cholesterol. |
| **Lumley, et al Female Model**^169^ | Lumley | 2002 | Cardiovascular health study | USA | 3393 | Cox Proportional Hazard Model | AF, Age, Creatinine, DM, History of Heart disease, Impaired fasting glucose, LVH, SBP, Sex, 15 ft walking time. |
| **Lumley, et al Combined Model**^169^ | Lumley | 2002 | Cardiovascular health study | USA | 5888 | Cox Proportional Hazard Model | AF, Age, Creatinine, DM, History of Heart disease, Impaired fasting glucose, LVH, SBP, Sex, 15 ft walking time. |
| **SPoRT Female Model**^170^ | Manuel | 2015 | Canadian Community Health Surveys | Canada | 44776 | Cox Proportional Hazard Model | Age, Alcohol, DM, Fruit and vegetable serving, Heart disease, Hypertension, Physical activity, Smoking, Stress. |
| **SpoRT Male Model**^170^ | Manuel | 2015 | Canadian Community Health Surveys | Canada | 44776 | Cox Proportional Hazard Model | Age, Alcohol, DM, Fruit and vegetable serving, Heart disease, Hypertension, Physical activity, Smoking, Stress. |
| **Framingham-Regicor Function Full Model Female**  ^171^ | Marrugat | 2014 | FRESCO Cohort Soutern Europe | Spain | 64824 | Cox Proportional Hazard Model | Age, anti-hypertensive medication use, BMI, Diabetes, HDL cholesterol, SBP, Smoking, Total Cholesterol. |
| **Framingham-Regicor Function Full Model Male**  ^171^ | Marrugat | 2014 | FRESCO Cohort Soutern Europe | Spain | 64824 | Cox Proportional Hazard Model | Age, anti-hypertensive medication use, BMI, Diabetes, HDL cholesterol, SBP, Smoking, Total Cholesterol. |
| **EUROSTROKE**^172^ | Moons | 2002 | (i) Kuopio Ischaemic Heart Disease Risk Factor Study  (ii) Rotterdam Study (iii) Caerphilly Heart Disease Study | (i) Finland (ii) NL (iii) UK | (i) 698 (ii) 148 (iii) 131 | Cox Proportional Hazard Model | Age, Current smoking, DBP, DM, Fibrinogen level, History of hypertension, History of stroke. |
| **FINRISK Lay-men  Female Model**^20^ | Qiao | 2012 | Finrisk Cohort | Finland | 16065 | Cox Proportional Hazard Model | Age, anti-hypertensive medication use, BMI, DM, fruit and vegetable consumption, happily married, incapable walking 500 meters, regular physical activity, Smoking. |
| **FINRISK Lay-men  Male Model**^20^ | Qiao | 2012 | Finrisk Cohort | Finland | 16065 | Cox Proportional Hazard Model | Age, anti-hypertensive medication use, BMI, DM, fruit and vegetable consumption, happily married, incapable walking 500 meters, regular physical activity, Smoking. |
| **CHINA-PAR**  **10-year Female Model**^19^ | Xing | 2019 | (i) China Multicenter Collaborative Study of Cardiovascular Epidemiology (CHINA MUCA)  (ii) International Collaborative Study of Cardiovascular Disease Asia | China | 21320 | Cox Proportional Hazard Model | Age, DM, Geographic region, HDL-Cholesterol, Smoking, Total Cholesterol, Urbanization, Untreated SBP, Waist circumference. |
| **CHINA-PAR**  **10-year Male Model**^19^ | Xing | 2019 | (i) China Multicenter Collaborative Study of Cardiovascular Epidemiology (CHINA MUCA)  (ii) International Collaborative Study of Cardiovascular Disease Asia | China | 21320 | Cox Proportional Hazard Model | Age, DM, Geographic region, HDL-Cholesterol, Parental History of Stroke, Smoking, Total Cholesterol, Urbanization, Untreated SBP. |
| **CHINA-PAR**  **Life-time Female Model**^19^ | Xing | 2019 | (i) China Multicenter Collaborative Study of Cardiovascular Epidemiology (CHINA MUCA)  (ii) International Collaborative Study of Cardiovascular Disease Asia | China | 21320 | Cox Proportional Hazard Model | Age, DM, Geographic region, HDL-Cholesterol, Smoking, Total Cholesterol, Urbanization, Untreated SBP, Waist circumference. |
| **CHINA-PAR**  **Life-time Male Model**^19^ | Xing | 2019 | (i) China Multicenter Collaborative Study of Cardiovascular Epidemiology (CHINA MUCA)  (ii) International Collaborative Study of Cardiovascular Disease Asia | China | 21320 | Cox Proportional Hazard Model | Age, DM, Geographic region, HDL-Cholesterol, Parental History of Stroke, Smoking, Total Cholesterol, Urbanization, Untreated SBP. |
| **Yao, et al.**^173^ | Yao | 2020 | China Health and Retirement Longitudinal Survey | China | 16557 | Logistic Regression Model | Age, DM, Heart disease, Hypertension, Smoking. |
| **Yatsuya 2013 Model**^174^ | Yatsuya | 2013 | Japanese public health center bases prospective cohort II | Japan | 15672 | Cox Proportional Hazard Model | Age, anti-hypertensive medicine use, BMI, DBP, DM, SBP, Sex, Smoking. |
| **Yatsuya 2016 Model**^175^ | Yatsuya | 2013 | Japanese public health center bases prospective cohort II | Japan | 15672 | Cox Proportional Hazard Model | Age, Anti-hypertensive medicine use, DM, HDL, SBP, Sex, Smoking. |
| **Zhang, et al Ischemic Model**^176^ | Zhang | 2005 | Chinese Steel worker cohort | China | 4400 | Cox Proportional Hazard Model | Age, SBP, Smoking, Total Cholesterol. |
| **Zhang, et al Hemorrhagic Model**^176^ | Zhang | 2005 | Chinese Steel worker cohort | China | 4400 | Cox Proportional Hazard Model | Age, DBP, SBP, Total Cholesterol. |
| **Dementia** | | | | | | | |
| **ANU-ADRI**^15^ | Anstey | 2013 | NA | NA | NA | Systematic review of the literature | Age, Alcohol, BMI, Cognitive Activity, DM, Education, Fish intake, High Cholesterol, Pesticide exposure, Physical activity, Smoking, Social Engagement, Symptoms of depression, TBI. |
| **CogDRisk**^177^ | Anstey | 2022 | (i) Rush Memory and Aging Project  (ii) The Health and Retirement Study - Aging, Demographs and Memory Study (iii) The Cardiovascular Health Study | USA | (i) 843  (ii) 421 (iii)3097 | Systematic review of the literature | AF, Age, BMI, Cognitive activity, Depression, DM, Education, Fish intake, History of stroke, Hypertension, Insomnia, Loneliness, Physical activity, Smoking, TBI, Total Cholesterol. |
| **Brief Dementia Screening indicator**^22^ | Barnes | 2014 | (i) Cardiovascular Health Study  (ii) Framingham Heart Study  (iii) Health and Retirement Study  (iv) Sacramento Area Latino Study on Aging | USA | 20219 | Cox Proportional Hazard Model | Age, Assistance with finances or medication, BMI, Depressive symptoms, DM, Education, History of Stroke |
| **Late-life Dementia Risk Index**^178^ | Barnes | 2009 | Cardiovascular Health Study | USA | 3375 | Logistic Regression Model | Age, 3MS, DSST, BMI, MRI WHM, Internal Carotid Artery Thickness, History of CABG, Time spent putting shirt on, Alcohol consumption |
| **MADeN**^179^ | Downer | 2016 | H-EPESE Cohort | USA | 1739 | Cox Proportional Hazard Model | Age, Attending Community Events, cannot walk ½ mile, DM, Education, Feel the Blues, Friends to count on, iADL impaired, Pain, Sex. |
| **DemPoRT Male Model**^21^ | Fisher | 2017 | Canadian Community Health Surveys | Canadian | 32297 | Cox Proportional Hazard Model | Age, Alcohol use, BMI, COPD, DM, Education, Epilepsy, Ethnicity, Fruit and Vegetables Consumption, Heart disease, High BP, Immigrant, Juice consumption, Marital status, Mood disorders, Multilingualism, Neighborhood deprivation, Number of activities required help from others, Physical activity, Potato Consumption, Sense of Belonging, Sex, Smoking, Stress, Stroke, Self-rated health. |
| **Hisayama Study Dementia**^180^ | Honda | 2021 | Hisayama Study | Japan | 795 | Cox Proportional Hazard Model | Age, BMI, DM, Education, History of stroke, Hypertension, Sex, Smoking. |
| **CAIDE**^181^ | Kivipelto | 2006 | CAIDE | Finland | 2293 | Cox Proportional Hazard Model | Age, BMI, Cholesterol, Education, Physical Inactivity, SBP, Sex |
| **Framingham Heart Study Dementia**^182^ | Li | 2018 | Framingham Heart Cohort | USA | 2372 | Cox Proportional Hazard Model | Age, BMI, Cancer, DM, Ischemic Attack, Martial status, Stroke. |
| **Mitniski, et al Model**^183^ | Mitniski | 2006 | Gothenburg H70 | Germany | 392 | Multivariate classification model | Age, Aortic calcification, BMI, Calf pain when walking – ceases when halts, Chest pain when excited, Cholesterol, Defective ventricular conduction, Pulmonary congestion, Sex. |
| **Ohara, et al Model**^184^ | Ohara | 2011 | Hisayama Study | Japan | 532 | Cox Proportional Hazard Model | Age, Alcohol intake, anti-hypertensive medication use, BMI, Education, HbA1c, regular exercise, SBP, Sex, Smoking, Total Cholesterol. |
| **LIBRA**^14^ | Schiepers | 2018 | The Maastricht Aging Study | NL | 949 | Cox Proportional Hazard Model | Age, Alcohol, Cholesterol, Cognitive Function, CVD, Depression, DM, Education, Hypertension, Obesity, Physical inactivity, Renal dysfunction, Sex, Smoking. |
| **Stephan, et al Complete Model**^185^ | Stephan | 2015 | Three City Study | France | 1172 | Cox Proportional Hazard Model | Age, Alcohol use, Apoe e4, Benton Visual Retention Test, Diabetes Use, Digit Span, Education, Impairment of activities of daily living, MMSE, MRI Hippocampal volume, MRI Whole Brain Volume, MRI WMH, Sex. |
| **Dementia Risk Model Age  60-79**^186^ | Walters | 2016 | The Health Improvement Network (THIN) | UK | 930395 | Cox Proportional Hazard Model | Age, Alcohol problem, BMI, Current anti-depressant medication, Current anti-hypertensive medication, Current aspirin use, Current Depression, DM, History of Stroke, History of TIA, Local area deprivation Cohort, Sex, Smoking. |
| **Dementia Risk Model Age  80-95**^186^ | Walters | 2016 | The Health Improvement Network (THIN) | UK | 930395 | Cox Proportional Hazard Model | AF, Age, BMI, Current anti-depressant medication, Current Depression, DM, HDL Cholesterol, History of Alcohol Problem, History of Stroke, History of TIA, SBP, Sex, Smoking, Total Cholesterol. |
| **Late-life depression** | | | | | | | |
| **DRAT-up**^17^ | Cattelai | 2010 | (i) English Longitudinal Study of Ageing (ii) Invecchiare nel Chianti (iii) The Irish Study of Ageing | (I) UK (II) Italy (iii) Ierland | 24698 | Systematic review of the literature | Bereavement, Current depression, Disability, Sex, Sleep disorder, Widowed. |
| **MANTO RPM FLR**^187^ | Murri | 2022 | European Share Cohort | EU | 39439 | Linear Regression Model | Able to regularly buy groceries, Activities in political organizations, Activities in religious organizations, Age, Age of onset of affective disorder, Appetite, Current occupation status, Depression, Difficulties carrying over 5 kg, Difficulties in getting up from a chair, Difficulties in sitting for two hours, Difficulties in walking 100 m, Difficulties one flight of stairs, Difficulties taking medication, Difficulty using a map in a strange place, Dizziness, Educational or training course, Enjoyment, Entering a nursing home, Family responsibilities prevent from doing things, Faints or blackouts, Feeling left out of things, Feeling out of control, Feel full of energy, Feel of opportunities, Financial stability, Future looks good, Guilt, Hearing function, Help from outside household, Irritability, Loneliness, Look back on life with happiness, Looking forward to each day, Lung disease, Marital status, No difficulties, Number of grandchildren, Pain, Parkinson, Pessimism, Playing word or number games, Reading ability, Reading books, newspapers, Recent bereavement, Rural/urban residence, Self-perceived health, Sex, Sleep, Smoking, Sport or a social or other kind of club, Suicidality, Tearfulness, Unable to afford a medical visit, Unable to see doctor due to waiting times, Use of antihypertensive medication, Use of drugs for anxiety or depression, Use of drugs for chronic bronchitis, Use of drugs for joint pain, Use of drugs for stomach burns, Use of hypnotics, Visual function, Weight loss, Widowed. |

Abbreviations: 3ms = Modified Mini-Mental State Examination, AF = Atrial Fibrillation, BMI = Body Mass Index, CABG = Coronary Artery Bypass Grafting, CHD = Coronary Heart Disease, CKD = Chronic Kidney Disease, COPD = Chronic Obstructive Pulmonary Disease, CVD = Cardiovascular Disease, DBP = Diastolic Blood Pressure, DM = Diabetes Mellitus, DSST = Digit Symbol Substitution Test, DVT = Deep Vein Thrombosis, EU = European Union, FH = Family History (often in the context of medical conditions), HDL = High-Density Lipoprotein, iADL= Instrumental Activities of Daily Living, kg = kilograms, LDL = Low-Density Lipoprotein, LVH = Left Ventricular Hypertrophy, m = meters, MMSE, = Mini-Mental State Examination, MRI = Magnetic Resonance Imaging, NA = Not Applicable, NL = Netherlands, PE = Pulmonary Embolism, RA = Rheumatoid Arthritis, SBP = Systolic Blood Pressure, TIA = Transient Ischemic Attack, UK = United Kingdom, USA = United States of America, WMH = White Matter Hyperintensities

**Table S5:** Selection reason meta-analyses for model development

| First Author | Year of publication | Risk factor | Risk factor categories | Reason for inclusion |
| --- | --- | --- | --- | --- |
| **Stroke** | | | | |
| Zhang^156^ | 2014 | Alcohol | Low, moderate, high alcohol consumption | Most recent MA, describing effect size as RR, largest sample size, no evidence of PB. No other MA with lower level of ROB or with unclear definitions. |
| Wang^43^ | 2017 | Hypertension | Hypertension | Most recent MA describing effect size as RR, largest sample size. PB not described.  No other MA with lower ROB or lower heterogeneity. |
| Huang^45^ | 2014 | Hypertension | Pre-hypertension | Most recent MA describing effect size as RR, largest sample size. ROB, and PB not described. No other MA with ≥ 20% larger population or lower heterogeneity. |
| Wang^158^ | 2022 | BMI | Obesity | Most recent MA describing effect size as RR, largest sample size. PB not described. No other MA with lower ROB. |
| Wang^158^ | 2022 | BMI | Overweight | Most recent MA describing effect size as RR, largest sample size. PB not described. No other with lower ROB or lower heterogeneity |
| Wang^158^ | 2022 | BMI | Underweight | Only MA providing an effect size as RR for this disease-risk factor relationship. |
| Shi ^63^ | 2021 | Diabetes | Diabetes | MA with >20% larger population as compared to most recent, describing effect size as RR, no evidence of PB. No other meta-analysis with lower ROB. |
| Shi ^63^ | 2021 | Diabetes | Pre-diabetes | MA with >20% larger population as compared to most recent, describing effect size as RR, no evidence of PB. No other meta-analysis with lower heterogeneity, or lower ROB, |
| Peters^69^ | 2016 | Cholesterol | Total cholesterol | MA with >20% larger population as compared to most recent, describing effect size as RR, no evidence of PB. No other meta-analysis with lower heterogeneity, or lower ROB. |
| Yuan^36^ | 2022 | Cholesterol | LDL | Only MA providing an effect size as RR for this disease-risk factor relationship. |
| Eurelings^76^ | 2018 | Depressive symptoms | Depressive symptoms | No MA available with effect sizes described as RR or HR. Only MA providing an effect size as OR for this disease-risk factor relationship. |
| Hu^77^ | 2014 | Diet | Vegetable consumption | MA with >20% larger population as compared to most recent, describing effect size as RR, PB not described. No other meta-analysis with lower ROB or lower heterogeneity. |
| Bechthold^78^ | 2019 | Diet | Fruit consumption | MA with >20% larger population as compared to most recent, describing effect size as RR, PB not described. No other meta-analysis with lower ROB or lower heterogeneity. |
| Hu^79^ | 2023 | Diet | Whole grain consumption | MA with similar sample size as compared more recent MA, but with lower heterogeneity, similar ROB, and no publication bias. |
| Bechthold^78^ | 2019 | Diet | Diary consumption | Most recent MA describing effect size as RR, earlier MA with >20% larger population but with higher level of heterogeneity and higher ROB. PB not described. |
| Bechthold^78^ | 2019 | Diet | Fish intake | MA with >20% larger population as compared to most recent, describing effect size as RR, PB not described. |
| Papp^80^ | 2023 | Diet | Poultry consumption | Most recent MA describing effect size as RR, largest sample size, PB not described. |
| Mendes^81^ | 2023 | Diet | Legume consumption | Most recent MA describing effect size as RR, largest sample size. No evidence of PB. No other MA with lower heterogeneity or ROB. |
| Shao^82^ | 2016 | Diet | Nut consumption | MA with >20% larger population as compared to most recent, describing effect size as RR, no evidence of PB. No other MA with lower heterogeneity or ROB. |
| Bechthold^78^ | 2019 | Diet | Red meat consumption | Most recent MA, describing effect size as RR, largest sample size, PB not described. No other MA with lower heterogeneity. |
| Wang^83^ | 2022 | Diet | SSB consumption | Most recent MA describing effect size as RR, largest sample size. No evidence of PB. No other MA with lower heterogeneity or ROB. |
| Rossi^84^ | 2015 | Diet | Dietary glycemic load consumption | Most recent MA, describing effect size as RR, largest sample size. ROB and PB not described. No other MA with lower heterogeneity. |
| Strazzullo^85^ | 2009 | Diet | Salt consumption | Only MA providing an effect size as RR for this disease-risk factor relationship. |
| Kang^86^ | 2020 | Diet | Saturated fat consumption | Most recent MA, describing effect size as RR, largest sample size. No evidence of PB. No other MA with lower heterogeneity or lower ROB. |
| Zhang^87^ | 2016 | Diet | Protein consumption | Most recent MA, describing effect size as RR, largest sample size. No evidence of PB. No other MA with lower level ROB. |
| Khosravipour^92^ | 2021 | Hearing | Hearing loss | No MA available with effect sizes described as RR or HR. Only MA providing an effect size as OR for this disease-risk factor relationship. |
| Masson^96^ | 2015 | Kidney disease | eGFR <30 ml/min/1.73 m2 | Only MA providing an effect size as RR for this disease-risk factor relationship. |
| Masson^96^ | 2015 | Kidney disease | eGFR 30-60 ml/min/1.73 m2 | Only MA providing an effect size as RR for this disease-risk factor relationship. |
| Masson^96^ | 2015 | Kidney disease | eGFR 60-90 ml/min/1.73 m2 | Most recent MA, describing effect size as RR, largest sample size. PB not described. No other MA with lower ROB. |
| Lee^100^ | 2003 | Physical activity | High level of physical activity | MA with >20% larger population as compared to most recent, describing effect size as RR, no evidence of PB, ROB not described. |
| Lee^100^ | 2003 | Physical activity | Moderate level of physical activity | MA with >20% larger population as compared to most recent, describing effect size as RR, no evidence of PB, ROB not described. |
| Jike^114^ | 2018 | Sleep | Long sleep | MA with >20% larger population as compared to most recent, describing effect size as RR, no evidence of PB. No other MA with lower ROB. |
| He^115^ | 2017 | Sleep | Short sleep | MA with >20% larger population as compared to most recent, describing effect size as RR, no evidence of PB. No other MA with lower ROB. |
| Wu^110^ | 2023 | Sleep | Insomnia | Only MA providing an effect size as RR for this disease-risk factor relationship. |
| Peters^124^ | 2013 | Smoking | Current | MA with >20% larger population as compared to most recent, describing effect size as RR, no evidence of PB. ROB not performed, No other MA with lower heterogeneity. |
| Peters^124^ | 2013 | Smoking | Former | Most recent MA, describing effect size as RR, largest sample size, no evidence of PB. ROB and heterogeneity not described. |
| Valtorta^130^ | 2016 | Social relationship | Isolation/Loneliness | Only MA providing an effect size as RR for this disease-risk factor relationship. |
| Park^131^ | 2022 | Social relationship | Social network | Only MA providing an effect size as RR for this disease-risk factor relationship. |
| Booth^137^ | 2015 | Stress | Stress | No MA available with effect sizes described as RR. Only MA providing an effect size as HR for this disease-risk factor relationship. |
| **Dementia** | | | | |
| Xu^41^ | 2017 | Alcohol | Low and Moderate consumption | Most recent MA, describing effect size as RR, no evidence of PB. One other MA with a >20% larger sample size but with unclear cut-offs and no assessment of ROB, heterogeneity or PB. |
| Xu^41^ | 2017 | Alcohol | High alcohol consumption | Only MA providing an effect size as RR for this disease-risk factor relationship. |
| Ou^49^ | 2020 | Hypertension | Late-life hypertension | Only MA providing an effect size as RR for this disease-risk factor relationship. |
| Ou^49^ | 2020 | Hypertension | Midlife hypertension | Most recent MA, describing effect size as RR, largest sample size, no evidence of PB. No other MA with lower level of ROB. |
| Qu^57^ | 2020 | BMI | Midlife obesity | Most recent MA, describing effect size as RR, largest sample size, no evidence of PB. No other MA with lower level of heterogeneity or ROB. |
| Qu^57^ | 2020 | BMI | Late-life obesity | Most recent MA, describing effect size as RR, largest sample size, no evidence of PB. No other MA with lower level of heterogeneity or ROB. |
| Qu^57^ | 2020 | BMI | Midlife overweight | Most recent MA, describing effect size as RR, largest sample size, no evidence of PB. No other MA with lower heterogeneity or ROB. |
| Qu^57^ | 2020 | BMI | Late-life overweight | Most recent MA, describing effect size as RR, largest sample size, no evidence of PB. No other MA with lower heterogeneity or ROB. |
| Qu^57^ | 2020 | BMI | Midlife underweight | Only MA providing an effect size as RR for this disease-risk factor relationship. |
| Qu^57^ | 2020 | BMI | Late-life underweight | Most recent MA, describing effect size as RR, largest sample size, no evidence of PB. No other MA with lower level of heterogeneity or ROB. |
| Xue^66^ | 2019 | Diabetes | Diabetes | Only MA providing an effect size as RR for this disease-risk factor relationship. |
| Xue^66^ | 2019 | Diabetes | Pre-diabetes | Only MA providing an effect size as RR for this disease-risk factor relationship. |
| Zhu^72^ | 2022 | Cholesterol | Total cholesterol | Most recent MA, describing effect size as Rr, largest sample size, no evidence of PB. No other meta-analysis with lower heterogeneity or ROB. |
| Yates^75^ | 2016 | Cognitive activity | Cognitive activity | Only MA providing an effect size as RR for this disease-risk factor relationship. |
| Talebi^88^ | 2023 | Diet | Diary consumption | Only MA providing an effect size as RR for this disease-risk factor relationship. |
| Bakre^89^ | 2018 | Diet | Fish consumption | MA with >20% larger population as compared to most recent, no evidence of PB, ROB not described. No other MA with lower ROB. |
| Sun^91^ | 2022 | Diet | SSB intake | No MA available with effect sizes described as RR. Only MA providing an effect size as HR for this disease-risk factor relationship. |
| Zhu^90^ | 2021 | Diet | Saturated fat consumption | Most recent MA, describing effect size as RR, largest sample size, no evidence of PB. No other MA with lower ROB. |
| Liang^93^ | 2021 | Hearing | Hearing loss | Only MA providing an effect size as RR for this disease-risk factor relationship. |
| Kjaergaard^97^ | 2022 | Kidney disease | eGFR <30 ml/min/1.73 m2 | No MA available with effect sizes described as RR. Only MA providing an effect size as HR for this disease-risk factor relationship. |
| Kjaergaard^97^ | 2022 | Kidney disease | eGFR 30-60 ml/min/1.73 m2 | No MA available with effect sizes described as RR. Only MA providing an effect size as HR for this disease-risk factor relationship. |
| Kjaergaard^97^ | 2022 | Kidney disease | eGFR 60-90 ml/min/1.73 m2 | No MA available with effect sizes described as RR. Only MA providing an effect size as HR for this disease-risk factor relationship. |
| Yuan^98^ | 2023 | Pain | Pain | No MA available with effect sizes described as RR or HR. Only MA providing an effect size as OR for this disease-risk factor relationship. |
| Iso-Markku^103^ | 2022 | Physical activity | High level of physical activity | Most recent MA, describing effect size as RR, largest sample size, indicating for some publication bias. No other MA with lower level of heterogeneity or ROB. |
| Lee^105^ | 2018 | Physical activity | Moderate level of physical activity | No MA available with effect sizes described as RR or HR. Most recent MA, describing effect size as OR, largest sample size, no evidence of PB. No other MA with lower level of heterogeneity or ROB. |
| Sutin^107^ | 2023 | Meaning in life | Meaning in life | No MA available with effect sizes described as RR or HR. Most recent MA, describing effect size as OR, largest sample size, no evidence of PB, ROB not described. No other MA with lower heterogeneity. |
| Fan^118^ | 2019 | Sleep | Long sleep | No MA available with effect sizes described as RR. Only MA providing an effect size as HR for this disease-risk factor relationship. |
| Fan^118^ | 2019 | Sleep | Short sleep | No MA available with effect sizes described as RR. Only MA providing an effect size as HR for this disease-risk factor relationship. |
| Shi^120^ | 2018 | Sleep | Insomnia | Most recent MA, providing effect size as RR, largest sample size, no evidence of PB. No other MA with lower heterogeneity or ROB. |
| Shi^120^ | 2018 | Sleep | Sleep disturbance | Only MA providing an effect size as RR for this disease-risk factor relationship. |
| Zhong^128^ | 2015 | Smoking | Current | Most recent MA, providing effect size as RR, largest sample size, no evidence of PB. No other MA with lower heterogeneity or ROB. |
| Zhong^128^ | 2015 | Smoking | Former | Most recent MA, providing effect size as RR, largest sample size, no evidence of PB. No other MA with lower heterogeneity or ROB. |
| Wang^136^ | 2023 | Social relationship | Isolation/Loneliness | Most recent MA, providing effect size as RR, largest sample size, PB not described. No other MA with lower ROB and less applicable definitions. |
| Wang^136^ | 2023 | Social relationship | Social network | Most recent MA, providing effect size as RR, largest sample size, PB not described. No other MA with lower ROB and less applicable definitions. |
| Franks^138^ | 2021 | Stress | Stress | No MA available with effect sizes described as RR. Only MA providing an effect size as HR for this disease-risk factor relationship. |
| **Late-life depression** | | | | |
| Long^157^ | 2015 | Hypertension | Hypertension | Only MA providing an effect size as RR for this disease-risk factor relationship. |
| Valkanova^74^ | 2013 | Cholesterol | Total cholesterol | No MA available with effect sizes described as RR or HR. Only MA providing an effect size as OR for this disease-risk factor relationship. |
| Lawrence^94^ | 2020 | Hearing | Hearing loss | No MA available with effect sizes described as RR or HR. Only MA providing an effect size as OR for this disease-risk factor relationship. |
| Hill Almeida^122^ | 2022 | Sleep | Sleep disturbance | Most recent MA, describing effect size as RR, largest sample size, no evidence of PB. No other MA with lower heterogeneity or ROB |
| Lawrence^94^ | 2020 | Smoking | Current | No MA available with effect sizes described as RR or HR. Only MA providing an effect size as OR for this disease-risk factor relationship. |

Abbreviations. BMI: Body Mass Index, eGFR: Estimated Glomerular Filtration Rate, HR: Hazard Ratio, MA: Meta-Analysis, OR: Odds Ratio, PB: Publication Bias, ROB: Risk of Bias, RR: Relative Risk, and SSB: Sugar-Sweetened Beverage.

**Table S6:** Characteristics of the included meta-analysis included for weighted composite risk factor estimate calculation

| **First Author** | **Year of publication** | **Specific population** | **Risk factor definition** | **Reference definition** | **Sample size (N=)** | **Events (N=)** | **Included studies (N=)** | **ROB Tool** | **ROB** | **Hetero-geneity** | **PB analy-sis?** | **PB** |
| --- | --- | --- | --- | --- | --- | --- | --- | --- | --- | --- | --- | --- |
| **Stroke** | | | | | | | | | | | |  |
| Zhang^156^ | 2014 | No | Low Alcohol  <15g/day | No alcohol | 233689 | NA | 9 | NOS | All studies ≥6 | Q statistic: 0.034 | Yes | No evidence of publication bias |
| Zhang^156^ | 2014 | No | Moderate Alcohol  15-30 g/day | No alcohol | 414146 | NA | 14 | NOS | All studies ≥6 | Q statistic: 0.006 | Yes | No evidence of publication bias |
| Zhang^156^ | 2014 | No | High Alcohol  >30g/day | No alcohol | 414146 | NA | 14 | NOS | All studies ≥6 | Q statistic: 0.596 | No | No evidence of publication bias |
| Wang^43^ | 2017 | Chinese only | Hypertension  ≥140/90 mmHg | No hypertension | 279088 | 9927 | 8 | NOS | Median: 7 | I^2^ :59% | No | NA |
| Huang^45^ | 2014 | No | High prehypertension 130-139 / 85-89 mmHg | normotension <120/80 mmHg | 762393 | NA | 11 | NA | NA | I^2^ :10 % | No | NA |
| Huang^45^ | 2014 | No | Low prehypertension 120-129 / 80-84 | normotension <120/80 mmHg | 762393 | NA | 11 | NA | NA | I^2^ :10 % | No | NA |
| Wang^158^ | 2022 | No | Obesity  BMI ≥30kg/m^2^ | Normal weight  18.5-25 kg/m^2^ | 4121082 | NA | 20 | NOS | All studies ≥7 | I^2^ :99% | No | NA |
| Wang^158^ | 2022 | No | Overweight BMI >25-29.9 kg/m^2^ | Normal weight  18.5-25 kg/m^2^ | 3458041 | NA | 19 | NOS | All studies ≥7 | I^2^ :85% | No | NA |
| Wang^158^ | 2022 | No | Underweight BMI <18.5 kg/m^2^ | Normal weight  18.5-25 kg/m^2^ | 3917021 | NA | 13 | NOS | All studies ≥7 | I^2^ :89% | No | NA |
| Shi ^63^ | 2021 | No | Highest FPG | Lowest FPG | 2524770 | NA | 16 | NOS | All studies ≥6 | I^2^ :47% | Yes | Eggers: 0.80 |
| Shi^63^ | 2021 | Non diabetic people (FPG<126) | Highest FPG | Lowest FPG | 2537596 | NA | 15 | NOS | All studies ≥6 | I^2^ :11% | Yes | Eggers 0.37 |
| Peters^69^ | 2016 | Female only | Highest Total Cholesterol | Lowest Total Cholesterol | 786621 | NA | 15 | NOS | All studies ≥7 | I^2^ :41% | Yes | Eggers: 0.66 |
| Peters^69^ | 2016 | Male only | Highest Total Cholesterol | Lowest Total Cholesterol | 786621 | NA | 15 | NOS | All studies ≥7 | I^2^ :23% | Yes | Eggers: 0.66 |
| Yuan^36^ | 2022 | Chinese only | LDL cholesterol | NA | NA | NA | 3 | NOS | All studies ≥6 | I^2^ :76% | No | NA |
| Eurelings^76^ | 2018 | No | Presence of depressive symptoms | Absence of depressive symptoms | 23337 | NA | 10 | NOS | NA | I^2^ :0% | Yes | No publication bias |
| Hu^77^ | 2014 | No | Highest Vegetables consumption | Lowest Vegetables consumption | 932545 | 14803 | 16 | NOS | All studies ≥6 | I^2^ :40% | No | Eggers: 0.78 |
| Bechthold^78^ | 2019 | No | Highest Fruit consumption | Lowest Fruit consumption | 845806 | 30523 | 17 | Nutri-Grade | Moderate ROB | I^2^ :40% | No | NA |
| Hu^79^ | 2023 | No | Highest Whole grain consumption | Lowest whole grain consumption | 877897 | 43643 | 8 | NOS | Moderate ROB | I^2^ :17% | Yes | Eggers: 0.481 |
| Bechthold^78^ | 2019 | No | Highest Dairy consumption | Lowest Dairy consumption | 452216 | 16887 | 12 | Nutri-Grade | Moderate ROB | I^2^ :43% | No | NA |
| Bechthold^78^ | 2019 | No | Highest Fish consumption | Lowest Fish consumption | 409128 | 14360 | 20 | Nutri-Grade | Moderate ROB | I^2^ :37% | No | NA |
| Papp^80^ | 2023 | No | Highest Poultry  consumption | Lowest Poultry consumption | 1290356 | NA | 9 | ROBINS-I | Moderate-High ROB | I^2^ :44% | No | NA |
| Mendes^81^ | 2023 | No | Highest Legumes consumption | Lowest Legumes consumption | 770735 | 13590 | 9 | ROBINS-I | Moderate-High ROB | I^2^ :37% | Yes | No publication bias |
| Shao^82^ | 2016 | No | Highest Nut  consumption | Lowest Nut consumption | 671301 | NA | 11 | NOS | All studies 6 | I^2^ :0% | Yes | Eggers: 0.932 |
| Bechthold^78^ | 2019 | No | Highest Red Meat consumption | Lowest Red Meat consumption | 341767 | 10541 | 7 | Nutri-Grade | Moderate ROB | I^2^ :0% | No | NA |
| Wang^83^ | 2022 | No | Highest SSB  consumption | Lowest SSB  consumption | 341767 | 10541 | 7 | Nutri-Grade | Moderate ROB | I^2^ :0% | No | NA |
| Rossi^84^ | 2015 | No | Highest dietary glycemic load consumption | Lowest dietary glycemic load consumption | 242132 | 3255 | 10 | NA | NA | I^2^ :0% | No | NA |
| Strazzullo^85^ | 2009 | No | Highest Salt consumption | Lowest Salt consumption | 147129 | 5346 | 14 | Downs and Black Score | Range 12-18 | I^2^ :61% | Yes | Eggers: 0.26 |
| Kang^86^ | 2020 | No | Highest Saturated Fat consumption | Lowest Saturated Fat consumption | 598435 | NA | 14 | NOS | All studies ≥7 | I^2^ :38% | Yes | Eggers: 0.36 |
| Zhang^87^ | 2016 | No | Highest protein consumption | Lowest protein consumption | 528982 | NA | 12 | NOS | Low ROB | I^2^ :67% | Yes | Eggers 0.106 |
| Khosravipour^92^ | 2021 | No | Presence of hearing loss | No hearing loss | 5014271 | NA | 8 | NOS | All studies ≥6 | I^2^ :41% | Yes | Eggers: 0.120 |
| Masson^96^ | 2015 | No | eGFR <30 ml/min/1.73 m2 | eGFR >90 ml/min/1.73 m2 | 1743690 | NA | 15 | NOS | NA | I2:72% | No | NA |
| Masson^96^ | 2015 | No | eGFR 30-60 ml/min/1.73 m2 | eGFR >90 ml/min/1.73 m2 | 2100733 | NA | 52 | NOS | NA | I2:72% | No | NA |
| Masson^96^ | 2015 | No | eGFR 60-90 ml/min/1.73 m2 | eGFR >90 ml/min/1.73 m2 | 861526 | NA | 15 | NOS | NA | I^2^ :72% | No | NA |
| Lee^100^ | 2003 | No | High level physical activity | Low level physical activity | 357791 | 4274 | 23 | NA | NA | I^2^ :78% | Yes | No publication bias |
| Lee^100^ | 2003 | No | Moderate level physical activity | Low level physical activity | 357791 | 4274 | 23 | NA | NA | I^2^ :78% | Yes | No publication bias |
| Jike^114^ | 2018 | No | Long sleep  (> 8 hours) | Normal sleep | 542218 | NA | 14 | NOS | All studies ≥6 | I^2^ :71% | Yes | No significant publication bias |
| He^115^ | 2017 | No | Short sleep  (<6 hours) | Normal sleep  (7 hours) | 528653 | NA | 12 | NOS | All studies ≥6 | I^2^ :49% | Yes | Eggers: 0.48 |
| Wu^110^ | 2023 | No | Insomnia | No insomnia | 23312 | NA | 2 | AMSTAR | 5 | I^2^ :0% | Yes | Eggers: 0.650 |
| Peters^124^ | 2013 | Female only | Current Smoking | Never | 3817289 | 39042 | 16 | NA | NA | I^2^ :3% | Yes | No significant publication bias |
| Peters^124^ | 2013 | Male only | Current Smoking | Never | 3817289 | 39042 | 16 | NA | NA | I^2^ :3% | Yes | No significant publication bias |
| Peters^124^ | 2013 | Female only | Former Smoking | Never | 3534330 | 36449 | 13 | NA | NA | NA | Yes | No significant publication bias |
| Peters^124^ | 2013 | Male only | Former Smoking | Never | 3534330 | 36449 | 13 | NA | NA | NA | Yes | No significant publication bias |
| Valtorta^130^ | 2016 | High income countries only | Social Isolation or loneliness | No social Isolation or loneliness | 105514 | 2577 | 9 | Healthcare Research and Quality Framework | Ranging low-high ROB | I^2^ :53% | No | NA |
| Park^131^ | 2022 | No | Having a larger social network | Not having a larger social network | 23576 | NA | 3 | ROBINS-I | Low Moderate ROB | I^2^ :21% | No | NA |
| Booth^137^ | 2015 | No | Exposed to general stress, work stress or stressfull life event | Not exposed to stress | 146859 | NA | 14 | NOS | All studies ≥5 | I^2^ :82% | Yes | Eggers: 0.0 |
| **Dementia** | | | | | | | | | | | | |
| Xu^41^ | 2017 | No | > 3 standard drinks/day | No alcohol | 7250 | NA | 4 | NOS | All studies ≥8 | NA | Yes | Significant publication bias, trim and fill performed |
| Xu^41^ | 2017 | No | 1-3 standard drinks/day | No alcohol | 7250 | NA | 4 | NOS | All studies ≥8 | NA | Yes | Significant publication bias, trim and fill performed |
| Xu^41^ | 2017 | No | >1 per week,  <1 standard drinks/day | No alcohol | 7250 | NA | 4 | NOS | All studies ≥8 | NA | Yes | Significant publication bias, trim and fill performed |
| Ou^49^ | 2020 | No | Midlife (<65) Hypertension | Normotension | 1320441 | 24717 | 9 | NOS | Moderate ROB | I^2^ :89% | No | NA |
| Ou^49^ | 2020 | No | Late-life (≥65) Hypertension | Normotension | 57907 | 4793 | 23 | NOS | Moderate ROB | I^2^ :32% | Yes | No evidence of publication bias |
| Qu^57^ | 2020 | No | Midlife (<65) Obesity | Normal weight | 41656 | NA | 8 | NOS | All studies ≥6 | I^2^ :44% | Yes | No significant publication bias |
| Qu^57^ | 2020 | No | Late-life (≥65) Obesity | Normal weight | 46472 | NA | 10 | NOS | All studies ≥6 | I^2^ :18% | Yes | No significant publication bias |
| Qu^57^ | 2020 | No | Midlife (<65) Overweight | Normal weight | 53690 | NA | 9 | NOS | All studies ≥6 | I^2^ :73% | Yes | No significant publication bias |
| Qu^57^ | 2020 | No | Late-life (≥65) Overweight | Normal weight | 32079 | NA | 8 | NOS | All studies ≥6 | I^2^ :38% | Yes | No significant publication bias |
| Qu^57^ | 2020 | No | Midlife (<65) Underweight | Normal weight | 33992 | NA | 6 | NOS | All studies ≥6 | I^2^ :0% | Yes | No significant publication bias |
| Qu^57^ | 2020 | No | Late-life (≥65) Underweight | Normal weight | 24556 | NA | 6 | NOS | All studies ≥6 | I^2^ :0% | Yes | No significant publication bias |
| Xue^66^ | 2019 | No | Fasting Plasma Glucose ≥7mmol/L | Fasting Plasma Glucose <7mmol/L | 2956619 | NA | 10 | NOS | Acceptable quality | I^2^ :16% | Yes | No publication bias |
| Xue^66^ | 2019 | No | Fasting Plasma Glucose ≥5.6mmol/L | Fasting Plasma Glucose <5.6mmol/L | 2956619 | NA | 10 | NOS | Acceptable quality | I^2^ :0% | Yes | No publication bias |
| Zhu^72^ | 2022 | No | Elevated Total Cholesterol | Not elevated total cholesterol | 153690 | 5638 | 9 | NOS | Low-moderate ROB | I^2^ :22% | Yes | Eggers: 0.824 |
| Yates^75^ | 2016 | No | Leisure time cognitive activity | No leisure time cognitive activity | 1932 | NA | 3 | CASP | NA | I^2^: 30-60% | No | NA |
| Talebi^88^ | 2023 | No | Highest Dairy intake | Lowest Dairy intake | 2757 | 693 | 3 | GRADE | Serious ROB | I^2^ :0% | Yes | No publication bias |
| Bakre^89^ | 2018 | No | More fish consumption | Less fish consumption | 40668 | 3139 | 9 | NA | NA | I^2^ :0% | Yes | Eggers: 0.597 |
| Sun^91^ | 2022 | No | Higher SSB intake | Lower SSB intake | 5660 | NA | 2 | NOS | All studies 9 | I^2^: 0% | No | NA |
| Zhu^90^ | 2021 | No | Highest Saturated Fat Intake | Lowest Saturated Fat Intake | 7395 | 314 | 2 | NOS | All studies 8 | I^2^:47% | Yes | No significant publication bias |
| Liang^93^ | 2021 | No | Hearing loss | No hearing loss | 725847 | NA | 12 | NOS | All studies ≥7 | I^2^ :86% | Yes | Potential publication bias, trim and fill performed |
| Kjaergaard^97^ | 2022 | No | eGFR <30 ml/min/1.73 m2 | eGFR >90 ml/min/1.73 m2 | 468699 | NA | 2 | NA | NA | I^2^ :40% | No | NA |
| Kjaergaard^97^ | 2022 | No | eGFR 30-60 ml/min/1.73 m2 | eGFR >90 ml/min/1.73 m2 | 468699 | NA | 2 | NA | NA | I^2^ :85% | No | NA |
| Kjaergaard^97^ | 2022 | No | eGFR 60-90 ml/min/1.73 m2 | eGFR >90 ml/min/1.73 m2 | 468699 | NA | 2 | NA | NA | I^2^ :0% | No | NA |
| Yuan ^98^ | 2023 | No | Prevalence of pain | No pain | Total: 1122503 | NA | 24 | NOS | All studies ≥7 | I^2^ :81% | No | NA |
| Iso-Markku^103^ | 2022 | No | Highest physical activity | Lowest physical activity | 257983 | 28120 | 49 | NOS | Ranging Low-High ROB | I^2^ :69% | Yes | Some Publication Bias |
| Lee^105^ | 2018 | No | Moderate amount of physical activity (>1 hours/week &>2 times/week) | Inactivity (<1 hour/week) | 75447 | NA | 21 | NOS | NA | I^2^ :47% | No | No Publication Bias |
| Sutin^107^ | 2023 | No | Meaning in life | No meaning in life | 202393 | 6899 | 8 | NA | NA | I^2^ :6% | No | NA |
| Fan^118^ | 2019 | No | Long sleep ≥9 hours | Normal sleep | 43412 | NA | 7 | NOS | All studies ≥7 | I^2^ :68% | Yes | Eggers: 0.21 |
| Fan^118^ | 2019 | No | Short Sleep  <5 range <7 | Normal sleep | 43412 | NA | 7 | NOS | All studies ≥7 | I^2^ :62% | Yes | Eggers: 0.21 |
| Shi^120^ | 2018 | No | Insomnia | No insomnia | 226167 | 23436 | 12 | NOS | All studies ≥5 | I^2^ :85% | Yes | No significant publication bias |
| Shi^120^ | 2018 | No | Sleep disturbance including breathing, length and insomnia | No sleep disturbance | 246786 | 25847 | 12 | NOS | All studies ≥5 | I^2^ :76% | Yes | No significant publication bias |
| Zhong^128^ | 2015 | No | Current Smoking | Never | 937392 | 14935 | 18 | NOS | NA | I^2^ :51% | Yes | No significant publication bias |
| Zhong^128^ | 2015 | No | Former Smoking | Never | 937392 | 14935 | 18 | NOS | NA | I^2^ :6% | Yes | No significant publication bias |
| Wang^136^ | 2023 | No | High social engagement | Low Social engagement | 83765 | NA | 10 | NOS | All studies ≥5 | I^2^ :78% | Yes | Eggers:0.001 |
| Wang^136^ | 2023 | No | Higher loneliness | Lower loneliness | 87264 | NA | 13 | NOS | All studies ≥5 | I^2^ :64% | No | NA |
| Franks^138^ | 2021 | No | Higher perceived stress | Lower perceived stress | 1882 | 203 | 2 | NOS | All studies ≥6 | I^2^ :0% | No | NA |
| **Late-Life Depression** | | | | | | | | | | | | |
| Long^157^ | 2015 | Aged ≥60 years | Hypertension  ≥140/90 mmHg | No hypertension | 9647 | 1690 | 5 | NOS | All studies ≥6 | I^2^ :65% | Yes | Eggers: 0.622 |
| Valkanova^74^ | 2013 | Aged ≥50 years | Dyslipidemia | Dyslipidemia | 17957 | NA | 10 | NA | NA | I^2^ :41% | Yes | Eggers: 0.72 |
| Lawrence^94^ | 2020 | Aged ≥60 years | Hearing loss | No Hearing loss | 147148 | NA | 35 | GRADE | Serious ROB | I^2^ :83% | Yes | Egger: 0.38 |
| Hill Almeida^122^ | 2022 | Aged ≥50 years | Self-reported sleep problem | Self-reported sleep problem | 54211 | NA | 14 | Cochrane ROB | Moderate-high ROB | I^2^ :16% | Yes | No obvious evidence of publication bias |
| Valkanova^74^ | 2013 | Aged ≥50 years | Smoking | Not smoking | 20120 | NA | 10 | NA | NA | I^2^ :67% | Yes | Egger: 0.85 |

**Abbreviations:** AMSTAR = AMeaSurement Tool to Assess systematic Reviews, CASP = Critical Appraisal Skills Program, eGFR = estimated Glomerular Filtration Rate, FPG = Fasting Plasma Glucose, GRADE = Grading of Recommendations Assessment, Development and Evaluation, kg/m^2 = kilograms per square meter (commonly used for Body Mass Index), LDL = Low-Density Lipoprotein, ml/min = milliliters per minute, mmHg = millimeters of mercury (a measurement of blood pressure), N = Number (often used in the context of sample size in studies), NA = Not Applicable, NOS = Newcastle-Ottawa Scale (a quality assessment tool for non-randomized studies), PB = Publication Bias, ROB = Risk Of Bias, ROBINS-I = Risk Of Bias In Non-randomized Studies of Interventions, SSB = Sugar-Sweetened Beverages.

**Table S7:** intermediate calculations

|  | **Stroke** | | | **Dementia** | | | **LLD** | | | **Stroke** | | **Dementia** | | **LLD** | |  | **Combined** | | | | | |
| --- | --- | --- | --- | --- | --- | --- | --- | --- | --- | --- | --- | --- | --- | --- | --- | --- | --- | --- | --- | --- | --- | --- |
| **Risk factor** | RR | 95%CI Lower | 95% Upper | RR | 95%CI Lower | 95% Upper | RR | 95%CI Lower | 95% Upper | Beta | Var | Beta | Var | Beta | Var | Total DALY | Beta | Var | RR | 95%CI Lower | 95% Upper | Nor. Beta |
| **Alcohol (ref never)** | | | | | | | | | | | | | | | | | | | | | | |
| > 3 drinks/day | 1.2 | 1.01 | 1.43 | 1.00 | 0.39 | 2.59 | NA | NA | NA | 0.1823 | 0.0115 | 0 | 0.3150 | NA | NA | 0.0665 | 0.1550 | 0.0154 | 1.17 | 0.92 | 1.94 | NS |
| 1-3 drinks/day | 1.01 | 0.93 | 1.09 | 0.58 | 0.38 | 0.90 | NA | NA | NA | 0.0100 | 0.0017 | -0.5447 | 0.0176 | NA | NA | 0.0665 | -0.0732 | 0.0016 | 0.93 | 0.85 | 1.01 | NS |
| <1 drinks/day | 0.85 | 0.75 | 0.95 | 0.75 | 0.51 | 1.11 | NA | NA | NA | -0.1625 | 0.0026 | -0.2877 | 0.0234 | NA | NA | 0.0665 | -0.1813 | 0.0024 | 0.83 | 0.74 | 0.93 | -20 |
| **Blood pressure (ref <120/80 mmHg)** | | | | | | | | | | | | | | | | | | | | | | |
| ≥140/90 ML | 2.68 | 2.2 | 3.26 | 1.2 | 1.06 | 1.36 | 1.16 | 0.91 | 1.42 | 0.9858 | 0.0731 | 0.1823 | 0.0059 | 0.1484 | 0.0169 | 0.0733 | 0.799 | 0.044 | 2.22 | 1.81 | 2.63 | 87 |
| ≥140/90 LL | 2.68 | 2.2 | 3.26 | 1.01 | 0.96 | 1.06 | 1.16 | 0.91 | 1.42 | 0.9858 | 0.0731 | 0.0100 | 0.0007 | 0.1484 | 0.0169 | 0.0733 | 0.775 | 0.044 | 2.17 | 1.76 | 2.58 | 85 |
| 130-139/ 85-89 | 1.95 | 1.73 | 2.21 | NA | NA | NA | NA | NA | NA | 0.6678 | 0.0150 | NA | NA | NA | NA | 0.0565 | 0.668 | 0.015 | 1.95 | 1.73 | 2.21 | 73 |
| 120-129 / 80-84 | 1.44 | 1.27 | 1.63 | NA | NA | NA | NA | NA | NA | 0.3646 | 0.0084 | NA | NA | NA | NA | 0.0565 | 0.365 | 0.008 | 1.44 | 1.27 | 1.63 | 40 |
| **BMI (ref 18.5-25 kg/m2)** | | | | | | | | | | | | | | | | | | | | | | |
| ≥30 ML | 1.47 | 1.02 | 2.11 | 1.45 | 1.19 | 1.78 | NA | NA | NA | 0.3853 | 0.0773 | 0.3716 | 0.0227 | NA | NA | 0.0665 | 0.3832 | 0.0564 | 1.47 | 1.00 | 1.93 | 42 |
| ≥30 LL | 1.47 | 1.02 | 2.11 | 0.78 | 0.7 | 0.86 | NA | NA | NA | 0.3853 | 0.0773 | -0.2485 | 0.0017 | NA | NA | 0.0665 | 0.2903 | 0.0559 | 1.34 | 0.87 | 1.80 | NS |
| 25-30 ML | 1.25 | 1.16 | 1.34 | 1.22 | 1.07 | 1.39 | NA | NA | NA | 0.2231 | 0.0021 | 0.1989 | 0.0067 | NA | NA | 0.0665 | 0.2195 | 0.0017 | 1.25 | 1.17 | 1.33 | 24 |
| 25-30 LL | 1.25 | 1.16 | 1.34 | 0.82 | 0.74 | 0.92 | NA | NA | NA | 0.2231 | 0.0021 | -0.1985 | 0.0021 | NA | NA | 0.0665 | 0.1600 | 0.0016 | 1.17 | 1.10 | 1.25 | 17 |
| <18.5 ML | 0.93 | 0.82 | 1.06 | 1.42 | 1.12 | 1.8 | NA | NA | NA | -0.0726 | 0.0037 | 0.3507 | 0.0301 | NA | NA | 0.0665 | -0.0092 | 0.0034 | 0.99 | 0.88 | 1.10 | NS |
| <18.5 LL | 0.93 | 0.82 | 1.06 | 1.23 | 1.03 | 1.48 | NA | NA | NA | -0.0726 | 0.0037 | 0.2070 | 0.0132 | NA | NA | 0.0665 | -0.0307 | 0.0030 | 0.97 | 0.86 | 1.08 | NS |
| **Blood Sugar (ref fasting plasma glucose < 100 mg/dL)** | | | | | | | | | | | | | | | | | | | | | | |
| >126 | 1.79 | 1.68 | 1.91 | 1.21 | 1.06 | 1.37 | NA | NA | NA | 0.5822 | 0.0034 | 0.1906 | 0.0063 | NA | NA | 0.0665 | 0.5235 | 0.0026 | 1.69 | 1.59 | 1.79 | 57 |
| 100-126 | 1.16 | 1.11 | 1.21 | 1.22 | 1.06 | 1.41 | NA | NA | NA | 0.1484 | 0.0007 | 0.1989 | 0.0080 | NA | NA | 0.0665 | 0.1560 | 0.0006 | 1.17 | 1.12 | 1.22 | 17 |
| **Cholesterol  (ref TC/LDL low)** | | | | | | | | | | | | | | | | | | | | | | |
| TC (F) | 0.99 | 0.87 | 1.12 | 1.13 | 1.04 | 1.22 | 1.08 | 0.99 | 1.38 | -0.0101 | 0.0041 | 0.1222 | 0.0021 | 0.0770 | 0.0099 | 0.0733 | 0.0160 | 0.0025 | 1.02 | 0.92 | 1.11 | NS |
| TC (M) | 1.14 | 1.03 | 1.27 | 1.13 | 1.04 | 1.22 | 1.08 | 0.99 | 1.38 | 0.1310 | 0.0037 | 0.1222 | 0.0021 | 0.0770 | 0.0099 | 0.0733 | 0.1248 | 0.0024 | 1.13 | 1.04 | 1.23 | 14 |
| LDL | 1.09 | 0.85 | 1.39 | NA | NA | NA | NA | NA | NA | NA | NA | NA | NA | NA | NA | NA | 0.0862 | NA | 1.09 | 0.85 | 1.39 | NS |
| **Cognitive activity (ref none)** | | | | | | | | | | | | | | | | | | | | | | |
| present | NA | NA | NA | 0.61 | 0.42 | 0.9 | NA | NA | NA | NA | NA | NA | NA | NA | NA | NA | -0.4943 | NA | 0.61 | 0.42 | 0.9 | -54 |
| **Depressive symptoms (ref not present)** | | | | | | | | | | | | | | | | | | | | | | |
| present | 1.36 | 1.13 | 1.51 | NA | NA | NA | NA | NA | NA | NA | NA | NA | NA | NA | NA | NA | 0.3075 | NA | 1.36 | 1.13 | 1.51 | 34 |
| **Diet (ref low)** | | | | | | | | | | | | | | | | | | | | | | |
| Vegetable intake | 0.86 | 0.79 | 0.93 | NA | NA | NA | NA | NA | NA | NA | NA | NA | NA | NA | NA | NA | -0.1508 | NA | 0.86 | 0.79 | 0.93 | -16 |
| Fruit intake | 0.83 | 0.77 | 0.89 | NA | NA | NA | NA | NA | NA | NA | NA | NA | NA | NA | NA | NA | -0.1863 | NA | 0.83 | 0.77 | 0.89 | -20 |
| Whole grain intake | 0.93 | 0.87 | 1 | NA | NA | NA | NA | NA | NA | NA | NA | NA | NA | NA | NA | NA | -0.0726 | NA | 0.93 | 0.87 | 1.00 | NS |
| Dairy intake | 0.96 | 0.9 | 1.01 | 0.72 | 0.58 | 0.89 | NA | NA | NA | -0.0408 | 0.0008 | -0.3285 | 0.0063 | NA | NA | 0.0665 | -0.0839 | 0.0007 | 0.92 | 0.87 | 0.97 | -9 |
| Fish intake | 0.95 | 0.89 | 1.01 | 0.8 | 0.74 | 0.87 | NA | NA | NA | -0.0513 | 0.0009 | -0.2231 | 0.0011 | NA | NA | 0.0665 | -0.0770 | 0.0007 | 0.93 | 0.87 | 0.98 | -8 |
| Poultry intake | 0.97 | 0.87 | 1.02 | NA | NA | NA | NA | NA | NA | NA | NA | NA | NA | NA | NA | NA | -0.0305 | NA | 0.97 | 0.87 | 1.02 | NS |
| Beans intake | 1 | 0.93 | 1.08 | NA | NA | NA | NA | NA | NA | NA | NA | NA | NA | NA | NA | NA | 0.0000 | NA | 1.00 | 0.93 | 1.08 | NS |
| Nuts intake | 0.88 | 0.8 | 0.97 | NA | NA | NA | NA | NA | NA | NA | NA | NA | NA | NA | NA | NA | -0.1278 | NA | 0.88 | 0.80 | 0.97 | -14 |
| Red meat intake | 1.16 | 1.08 | 1.25 | NA | NA | NA | NA | NA | NA | NA | NA | NA | NA | NA | NA | NA | 0.1484 | NA | 1.16 | 1.08 | 1.25 | 16 |
| SSB intake | 1.12 | 1.03 | 1.23 | 2.77 | 2.23 | 3.43 | NA | NA | NA | 0.1133 | 0.0026 | 1.0188 | 0.0937 | NA | NA | 0.0665 | 0.2490 | 0.0040 | 1.28 | 1.16 | 1.41 | 27 |
| Sweets intake | 1.23 | 1.07 | 1.47 | NA | NA | NA | NA | NA | NA | NA | NA | NA | NA | NA | NA | NA | 0.2070 | NA | 1.23 | 1.07 | 1.47 | 23 |
| Sodium intake | 1.23 | 1.06 | 1.43 | NA | NA | NA | NA | NA | NA | NA | NA | NA | NA | NA | NA | 0.0565 | 0.2070 | NA | 1.23 | 1.06 | 1.43 | 23 |
| Saturated fats intake | 0.87 | 0.78 | 0.96 | 1.37 | 0.7 | 2.69 | NA | NA | NA | -0.1393 | 0.0021 | 0.3148 | 0.2577 | NA | NA | 0.0665 | -0.0712 | 0.0073 | 0.93 | 0.76 | 1.10 | NS |
| Protein intake | 0.98 | 0.89 | 1.07 | NA | NA | NA | NA | NA | NA | NA | NA | NA | NA | NA | NA | NA | -0.0202 | NA | 0.98 | 0.89 | 1.07 | NS |
| **Hearing loss (ref no loss)** | | | | | | | | | | | | | | | | | | | | | | |
| Present | 1.33 | 1.18 | 1.49 | 1.59 | 1.37 | 1.86 | 1.47 | 1.31 | 1.65 | 0.2852 | 0.0063 | 0.4637 | 0.0156 | 0.3646 | 0.0063 | 0.0733 | 0.3168 | 0.0041 | 1.37 | 1.25 | 1.50 | 35 |
| **Kidney function (ref eGFR ≥90 ml/min/1.73 m2)** | | | | | | | | | | | | | | | | | | | | | | |
| eGFR <30 | 1.7 | 1.47 | 1.96 | 1.91 | 1.21 | 3.01 | NA | NA | NA | 0.5306 | 0.0156 | 0.6471 | 0.2108 | NA | NA | 0.0665 | 0.5481 | 0.0160 | 1.73 | 1.48 | 1.98 | 60 |
| eGFR 30-60 | 1.43 | 1.33 | 1.54 | 1.31 | 0.92 | 1.87 | NA | NA | NA | 0.3577 | 0.0029 | 0.2700 | 0.0587 | NA | NA | 0.0665 | 0.3445 | 0.0034 | 1.41 | 1.30 | 1.53 | 38 |
| eGFR 60-90 | 1.1 | 1.03 | 1.19 | 1.14 | 1.06 | 1.22 | NA | NA | NA | 0.0953 | 0.0017 | 0.1310 | 0.0017 | NA | NA | 0.0665 | 0.1007 | 0.0012 | 1.11 | 1.04 | 1.17 | 11 |
| **Pain (ref no pain)** | | | | | | | | | | | | | | | | | | | | | | |
| Present | NA | NA | NA | 1.26 | 1.18 | 1.35 | NA | NA | NA | NA | NA | NA | NA | NA | NA | NA | 0.2311 | NA | 1.26 | 1.18 | 1.35 | 25 |
| **Physical activity (ref low)** | | | | | | | | | | | | | | | | | | | | | | |
| Moderate activity | 0.8 | 0.74 | 0.86 | 0.80 | 0.77 | 0.84 | NA | NA | NA | -0.2231 | 0.0009 | -0.2614 | 0.0001 | NA | NA | 0.0665 | -0.2289 | 0.0007 | 0.80 | 0.74 | 0.85 | -25 |
| High activity | 0.73 | 0.67 | 0.79 | 0.77 | 0.75 | 0.79 | NA | NA | NA | -0.3147 | 0.0009 | -0.2614 | 0.0001 | NA | NA | 0.0665 | -0.3067 | 0.0007 | 0.74 | 0.68 | 0.79 | -34 |
| **Sense of belonging/ purpose in life (ref no purpose)** | | | | | | | | | | | | | | | | | | | | | | |
| Present | NA | NA | NA | 0.76 | 0.72 | 0.79 | NA | NA | NA | NA | NA | NA | NA | NA | NA | NA | -0.2744 | NA | 0.76 | 0.72 | 0.79 | -30 |
| **Sleep (ref 6-8 hours)** | | | | | | | | | | | | | | | | | | | | | | |
| >8 hours | 1.46 | 1.26 | 1.69 | 1.77 | 1.32 | 2.37 | NA | NA | NA | 0.3784 | 0.0120 | 0.5710 | 0.0717 | NA | NA | 0.0665 | 0.4073 | 0.0103 | 1.50 | 1.30 | 1.70 | 44 |
| <6 hours | 1.1 | 0.97 | 1.24 | 1.2 | 0.91 | 1.59 | NA | NA | NA | 0.0953 | 0.0047 | 0.1823 | 0.0301 | NA | NA | 0.0665 | 0.1083 | 0.0041 | 1.11 | 0.99 | 1.24 | NS |
| insomnia | 1.55 | 1.39 | 1.75 | 1.17 | 0.95 | 1.43 | NA | NA | NA | 0.4383 | 0.0084 | 0.1570 | 0.0150 | NA | NA | 0.0665 | 0.3961 | 0.0064 | 1.49 | 1.33 | 1.64 | 43 |
| problem/disturbance | NA | NA | NA | 1.19 | 1.11 | 1.29 | 1.82 | 1.69 | 1.97 | NA | NA | 0.173953307 | 0.002108496 | 0.598836501 | 0.005102041 | 0.0168 | 0.3463 | 0.0016 | 1.41 | 1.34 | 1.49 | 38 |
| **Smoking  (ref never)** | | | | | | | | | | | | | | | | | | | | | | |
| Current (F) | 1.83 | 1.58 | 2.12 | 1.3 | 1.18 | 1.45 | 1.35 | 1.00 | 1.81 | 0.6043 | 0.0190 | 0.2624 | 0.0047 | 0.2776 | 0.0366 | 0.0733 | 0.5275 | 0.0117 | 1.69 | 1.48 | 1.91 | 58 |
| Current (M) | 1.67 | 1.49 | 1.88 | 1.3 | 1.18 | 1.45 | 1.35 | 1.00 | 1.81 | 0.5128 | 0.0099 | 0.2624 | 0.0047 | 0.2776 | 0.0366 | 0.0733 | 0.4570 | 0.0063 | 1.58 | 1.42 | 1.73 | 50 |
| Former (F) | 1.17 | 1.12 | 1.22 | 1.01 | 0.96 | 1.06 | NA | NA | NA | 0.1570 | 0.0007 | 0.0100 | 0.0007 | NA | NA | 0.0665 | 0.1350 | 0.0005 | 1.14 | 1.10 | 1.19 | 15 |
| Former (M) | 1.08 | 1.03 | 1.13 | 1.01 | 0.96 | 1.06 | NA | NA | NA | 0.0770 | 0.0007 | 0.0100 | 0.0007 | NA | NA | 0.0665 | 0.0669 | 0.0005 | 1.07 | 1.03 | 1.11 | 7 |
| **Social engagement (ref not lonely. small social network)** | | | | | | | | | | | | | | | | | | | | | | |
| Lonely | 1.32 | 1.04 | 1.68 | 1.42 | 1.26 | 1.6 | NA | NA | NA | 0.2776 | 0.0267 | 0.3507 | 0.0075 | NA | NA | 0.0665 | 0.2886 | 0.0194 | 1.33 | 1.06 | 1.61 | 32 |
| Large network | 0.77 | 0.57 | 1.04 | 0.81 | 0.74 | 0.89 | NA | NA | NA | -0.2614 | 0.0144 | -0.2107 | 0.0015 | NA | NA | 0.0665 | -0.2538 | 0.0104 | 0.78 | 0.58 | 0.98 | -28 |
| **Stress (ref no perceived stress)** | | | | | | | | | | | | | | | | | | | | | | |
| Present | 1.33 | 1.17 | 1.50 | 1.44 | 1.07 | 1.95 | NA | NA | NA | 0.2852 | 0.0094 | 0.3646 | 0.0504 | NA | NA | 0.0665 | 0.2971 | 0.0079 | 1.35 | 1.17 | 1.52 | 32 |

**Legend**. Abbreviations: CI: Confidence interval, dL: deciliter, eGFR: estimated Glomerulus Filtration Rate, Kg = kilograms, LDL = Low Density Lipoprotein, LL: Late-life, mg: milligrams, ML: Midlife, mmHg: millimeters, Ref: Reference Hg: mercury, NA: Not Applicable, NS: Not Significant, TC: Total Cholesterol.
